# Human milk oligosaccharides, oral rotavirus vaccine seroconversion, and rotavirus gastroenteritis risk in a vaccinated birth cohort

**DOI:** 10.64898/2025.12.04.25341660

**Authors:** Rebecca J. Rubinstein, Nadja A. Vielot, Jessie Edwards, Yaoska Reyes, Fredman González, Christian Toval-Ruíz, Lester Gutiérrez, Slavica Mijatovic-Rustempasic, Samuel Vilchez, Robert S. Sandler, Lakshmanane Premkumar, Lars Bode, Sylvia Becker-Dreps, Filemón Bucardo

## Abstract

**Objective:** Rotavirus is the leading cause of diarrhea-related mortality in children worldwide and oral rotavirus vaccines (ORVs) have a lower effectiveness in low- and middle-income countries as compared to high-income countries. Novel strategies that leverage nature’s existing system of protection against rotavirus, such as from human milk, could guide future interventions to reduce global rotavirus burden. The goal of this study was to understand whether human milk oligosaccharides (HMOs) influence ORV response and the risk of rotavirus gastroenteritis in early life.

**Methods:** In a population-based cohort of 297 Nicaraguan infants followed weekly for 3 years, we assessed the relationships between HMO composition, IgA seroconversion after ORV, and subsequent rotavirus gastroenteritis incidence.

**Results:** Among all children, 17.5% experienced ≥ 1 symptomatic rotavirus gastroenteritis episode over 36 months of follow-up. Of the 297 children assessed through serology, only 34.7% seroconverted to the vaccine strain G1P[8]. Seroconverters experienced rotavirus gastroenteritis later and less frequently than non-seroconverters. Receiving milk with higher concentrations of 2’-fucosyllactose (2’FL) was associated with a greater likelihood of seroconversion to ORV. In contrast, receiving milk with several other fucosylated and sialylated HMOs was associated with decreased seroconversion to ORV. Additionally, receiving milk with higher concentrations of DFLNH (Difucosyllacto-N-hexaose) was associated with a decreased risk rotavirus gastroenteritis in the first 3 years of life, while receiving milk with higher concentrations of lacto-N-fucopentaose (LNFP-I) was associated with an increased risk.

**Conclusion:** While we found associations between levels of certain HMOs and either ORV seroconversion or rotavirus gastroenteritis risk, the relationships between milk composition, vaccine response, and infection risk are complex and highlight the need for careful safety assessment of HMO supplementation in settings with high rotavirus burden.

**Article Summary:** In a Nicaraguan birth cohort with low rotavirus vaccine immunogenicity, we evaluated the role of human milk oligosaccharides (HMOs) on rotavirus vaccine seroconversion and rotavirus gastroenteritis risk over the first 3 years of life.

**What’s Known on This Subject:** Rotavirus is the leading cause of diarrhea-related mortality in under-5 children worldwide, partly due to poor vaccine immunogenicity in low-and-middle-income countries. Maternal and child genetic secretor status is associated with vaccine response, gastroenteritis susceptibility, and HMO composition in human milk.

**What This Study Adds:** This study revealed that the concentrations of several fucosylated and sialylated HMOs had both positive and negative associations with rotavirus vaccine seroconversion and rotavirus gastroenteritis risk in children. Careful consideration must be taken prior to the use of HMOs as supplements.

## Introduction

Rotavirus remains the leading cause of diarrhea-related morbidity and mortality in children under 5 years worldwide, despite the global roll-out of licensed vaccines^1^. Performance of oral rotavirus vaccines (ORV) is lower in low- and middle-income countries (LMICs) (30%-88% effectiveness) than in high-income countries (95% effectiveness)^2,3^.

Rotavirus is spread through the “fecal-oral” route. However, in addition to environmental factors, genetic factors, including “secretor status” are associated with disease risk^4^. “Secretor” individuals have functional fucosyltransferase-2 (FUT2) enzymes that synthesize α-1-2 fucosylated glycans. These glycans are displayed on the intestinal epithelium in secretor individuals and have been shown to facilitate rotavirus attachment, making secretors more susceptible to rotavirus gastroenteritis^5^. In addition, FUT2 and other enzymes shape the distribution of human milk oligosaccharides (HMOs) which are secreted into the milk of lactating women.^6,7^ HMOs are a diverse group of glycans in human milk that are divided into three categories based on their chemical structures—fucosylated, neutral non-fucosylated, and sialylated. Fucosylated HMOs secreted into the milk of secretor women may serve as soluble attachment factors (“decoy receptors”) for rotavirus, preventing its attachment to the child’epithelium^4^. These same HMOs may prevent seroconversion by oral rotavirus vaccines, by reducing the live attenuated vaccine virus replication in the child’s intestines^8,9,6,10^. However, these relationships are complex—for example, HMOs paradoxically increase the infectivity of neonatal G10P[11] rotavirus^11^. HMOs have additional longer-term effects. HMOs are metabolized by gut commensal bacteria^11^, providing a competitive advantage to specific bacteria. They also shape the developing immune system^7,12^, which may modulate acute gastroenteritis (AGE) risk. Some commercially-available infant formulas already contain several HMOs, including 2’ fucosyllactose (2’FL), while clinical trials are testing additional HMOs as formula enhancers or as direct dietary supplements to formula-fed and breastfed children^13^. However, potential opposing effects of HMOs on vaccine response and rotavirus AGE risk must be clarified before the broader use of HMOs in infant formula and as supplements.

Nicaragua is a Central American LMIC with high childhood diarrheal disease burden^4,14–16^ and low estimated ORV effectiveness of 46%^17^. In our Nicaraguan birth cohort fully vaccinated with the 2-dose monovalent G1P[8] ORV (Rotarix, RV1©, GlaxoSmithKline), secretor children experienced more rotavirus AGE than non-secretors over the first three years of life^4^. Here, we investigate the relationship between maternal HMO concentrations, maternal and child secretor status, RV1 seroconversion and rotavirus AGE. These data could support supplementing breastfed and formula-fed children’s diets with HMOs as well as generate hypotheses for the biological pathways between maternal secretor status, HMOs, ORV performance and rotavirus susceptibility in children.

## Methods

### Cohort recruitment

The Sapovirus Acute GastroEnteritis (SAGE) study was a population-based birth cohort of 444 mother-infant dyads in a socioeconomically-diverse area of León, Nicaragua. All infant participants were enrolled within the first 10 days of life between June 2017 and July 2018 and vaccinated with 2 doses of RV1 at 2 and 4 months of age at local public health posts^14^. We collected information on sociodemographic, family, and household factors at baseline. Through weekly household visits, we assessed participants for AGE and reflexively collected stools for AGE episodes reported over 36 months. We defined incident AGE episodes as vomiting or diarrhea (increase in stool frequency to 3 stools per 24-hour period or a substantial change in stool consistency) following 3 days without symptoms^14^. While the original study was focused on sapovirus, we also tested AGE stools for rotavirus. Each week, we assessed for continued breastfeeding and complementary food introduction. Each month, lactating mothers provided self-expressed milk samples from one breast, at least one hour after not breastfeeding from that breast^14^. Mothers provided written informed consent for participation and biobanking of samples for future research. The Ethical Committee for Biomedical Research at the Universidad Nacional Autónoma de Nicaragua—León (UNAN-León) (Acta #2-2017) and the Institutional Review Board at the University of North Carolina at Chapel Hill (protocol #16-2079)^15^ provided ethical approval.

### Rotavirus detection, genotyping and rotavirus AGE definition

AGE stools (N=1505) were tested for rotavirus using reverse-transcriptase quantitative polymerase chain reaction (qRT-PCR)^4,18–20^. Sequencing and genotyping of G-types G1, G2, G3, equine-like G3, G4, G9, and G12 and P-types P[8], P[6], and P[4] was performed as described previously^4,21^. Sequencing of Rotarix non-structural-protein-2 gene distinguished “vaccine-derived” from wild-type rotavirus infections^4,22^, hereafter referred to as “rotavirus AGE”.

### Seroconversion status

We quantified Rotarix-specific IgA titers in infants’ serum approximately 2 weeks before the initial 2-month RV1 dose and approximately 1 month after the RV1 4-month dose using a modified enzyme-linked immunosorbent assay (ELISA) from Bucardo et al., (2015)^23^. We coated 96-well microtiter plates (Greiner Bio-One, Kremsmünster, Austria) with Wa (tissue-culture adapted) rotavirus A G1P[8] strain “VR-2018” from American Type Culture Collection^24^. After blocking, we added 100 µl of serially diluted plasma (1:20-1:640) and incubated plates at 37 °C for 1 hr. We detected IgA using horseradish peroxidase-conjugated goat anti-human IgA (Invitrogen A18781, Invitrogen, ThermoFisher Scientific, Waltham, MA) and 1-Step™ Ultra TMB-ELISA Substrate (Thermo Fisher Scientific, Stockholm, Sweden). We quantified titers using the reciprocal of the highest plasma dilution with Optical Density_450_ ≥ 0.100. Plasma IgA titers ≥1:80 were considered positive for Rotarix-specific IgA, and we defined seroconversion as a ≥ 4-fold increase in IgA titers between pre- and post-vaccination serum^23^.

### HMO composition

Nineteen HMOs, known to comprise approximately 95% of all HMOs found in human milk by concentration, were identified and quantified (in µg/mL) in milk samples using high pressure liquid chromatography after fluorescent labeling (HPLC-FL), a technique developed by the Bode Laboratory^25^. We assessed HMOs at approximately 1 month postpartum to capture the largest numbers of mothers who would still be breastfeeding their infants^26^. By one month postpartum, milk is considered mature, and HMO levels are less likely to fluctuate substantially as in the early postpartum period^27^.

### HBGA secretor and Lewis phenotypes

We considered mothers with 2’FL concentration ≥ 100 nmol/mL secretors, as FUT2-dependent 2’FL synthesis occurs only in secretors^28^. We ascertained child secretor status from baseline saliva samples using an in-house lectin-based ELISA targeting FUT-2-derived Fuc alpha 1-2Gal-R antigens^15^ and child Lewis status (reflecting FUT3 expression) using an ELISA targeting Lewis A and B antigens^29^.

### Statistical analysis

#### Baseline cohort characteristics

We described baseline medians (interquartile range [IQR]) and frequencies of continuous and categorical cohort characteristics, respectively. Children were “exclusively breastfed” until the first week when a mother reported feeding her child formula, liquids, or complementary foods. We defined weaning as the final weekly visit in which the mother reported any breastfeeding over 36 months of surveillance.

#### Rotavirus AGE cumulative incidence

To estimate effects of seroconversion and pre-vaccination IgA titers on AGE, we calculated the cumulative incidence of rotavirus AGE from 1-month post-vaccination to 36 months of age among cohort children. We examined the incidence in seroconverters and non-seroconverters, and in children with pre-vaccination IgA titers ≥80. We confirmed children’s vaccination status through immunization cards or parental reports during weekly visits. We initiated follow-up on the day that we quantified post-vaccination titers in serum (approximately one month after the 2^nd^ RV1 dose) or 30 days after the recorded or imputed 2^nd^ vaccination date for participants not assessed for seroconversion. We used the inverse probability of treatment weighted (IPTW) Kaplan-Meier survival function to determine the time-to-first rotavirus AGE episode, under each exposure (Supplementary Information). We used a causal diagram to identify potential confounders of the relationships between seroconversion and rotavirus AGE and pre-vaccination IgA titer ≥80 and rotavirus AGE. We calculated IPTWs using binary logistic regression based on confounders comprising the minimally-sufficient adjustment set identified by DAGitty (dagitty.net)^30^.

#### Crude associations between HMO composition, RV1 seroconversion, and rotavirus AGE

For each HMO, we assessed the probability of RV1 seroconversion by tertile of HMO concentration (highest vs. lowest tertile, and intermediate vs. lowest tertile to understand possible dose-dependent relationships) with a log-binomial linear risk model generating relative risks (RRs) (95% CI). We also applied a Kaplan-Meier survival function to calculate the 36-month cumulative incidence of rotavirus AGE among children consuming the highest or intermediate vs. lowest tertile HMO concentrations. We then calculated the 36-month risk difference (RD) (95% CI) by subtracting the rotavirus AGE risk among children consuming the lowest concentrations from children consuming the highest and intermediate concentrations. All analyses were further stratified by child secretor status, classified as an effect measure modifier in the analysis. Secretors express glycans on the intestinal mucosa that serve as attachment factors for rotavirus, and therefore have a substantially higher rate of rotavirus AGE as compared to non-secretor children^4^.

#### Adjusted associations between HMO composition, RV1 seroconversion and rotavirus AGE

We used a causal diagram to identify potential confounders of the relationships between HMOs and RV1 seroconversion, and HMOs and rotavirus AGE (Fig S1), calculating IPTWs as described above. Confounders included cumulative weeks of exclusive breastfeeding at HMO measurement, child’s age (approximating lactation stage) at HMO measurement^31^, maternal age at child’s birth, number of child’s older siblings (approximating parity)^28^, child secretor and Lewis phenotype^7^, and household earthen flooring (approximating poverty)^16^. We also included HMOs moderately-to-highly-correlated (Pearson r 0.5-1) with the primary HMO exposure in IPTWs to address confounding by co-occurring HMOs (Fig S2-S3). IPTWs were calculated using multinomial logistic regression (Supplementary Information).

#### Associations between HMO composition, RV1 seroconversion, and rotavirus AGE upon censoring children after weaning

We also investigated whether associations between HMO concentration and rotavirus AGE incidence changed after censoring children upon weaning. While HMOs can have longer-term effects through shaping the gut microbiome, decoy-receptor activity could only occur if HMOs are present in the gastrointestinal tract^32^. To correct for potential selection bias introduced by this artificial censoring^33^, we applied inverse probability of censoring weights (IPCWs) based on time-invariant and time-varying variables in a minimally-sufficient adjustment set associated with both weaning and rotavirus AGE (Supplementary Information, Fig S4). In models additionally adjusted for baseline confounders, we weighted models by the product of the IPTWs and IPCWs.

## Results

### Cohort characteristics

Within the original SAGE cohort (n=444 mother-child dyads), we excluded 25 dyads who did not initiate breastfeeding, 10 dyads missing covariate data, and an additional 27 dyads who dropped out prior to 12 months of infant age. All infants within the remaining 382 dyads received both RV1 doses before 12 months age, but only the 297 dyads with complete infant serum sets available at the 1-, 5- and 12-month timepoints were assessed for seroconversion (Fig 1). Characteristics were similar for the 297 cohort children who were assessed for seroconversion and the 85 cohort children who were not assessed.

**Figure 1.**
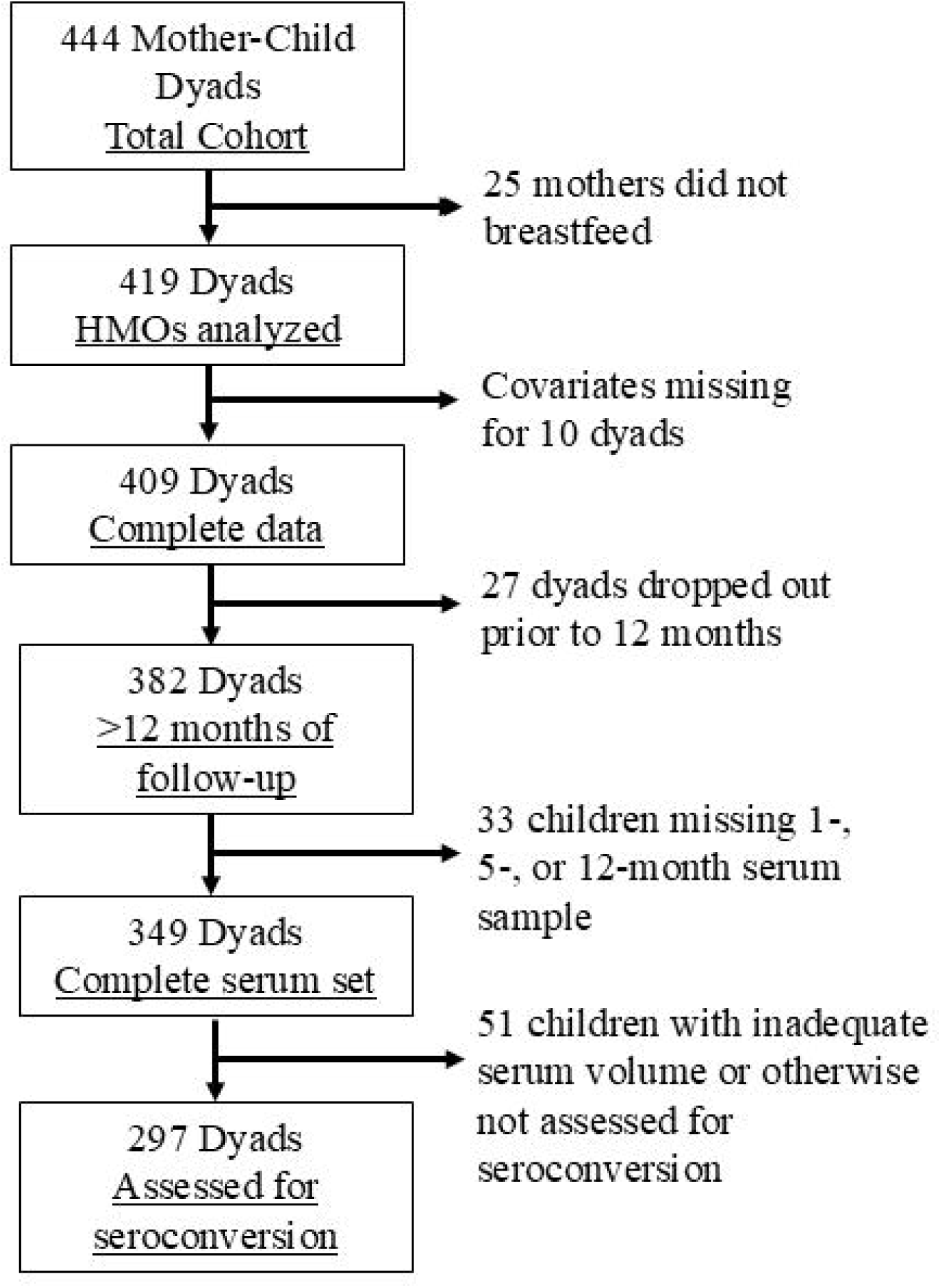
Flowchart for study inclusion.

Human milk analyzed for HMO concentrations was collected at a median of 1.3 months post-partum (range 0.2, 3.2). Duration of non-exclusive breastfeeding in the cohort was high, lasting a median of 22.8 months (interquartile range [IQR] 10.8, 32.1).

### Seroconversion and HBGA phenotypes

Of the 297 children assessed for serum IgA with pre- and post-vaccine paired samples, 34.7% seroconverted following the second dose of RV1. Secretors comprised 93.5% of mothers and 87.9% of children, and 76.1% of cohort children had the Lewis B phenotype, for which secretor status is a prerequisite. Neither maternal secretor status (*p=0.*77), child secretor status (*p=0.*35), nor child Lewis status (*p=0.*43) were associated with RV1 seroconversion (Table 1). Among participants with positive pre-vaccination IgA titers, 56/58 (96.6%) failed to seroconvert following vaccination. After removing these 58 participants, seroconversion in the cohort nonetheless remained low at 42.3% (101/239).

**Table 1:** Participant characteristics by monovalent rotavirus vaccine (RV1) seroconversion status.

| Characteristics | Frequency (%) or median (IQR<sup>a</sup>) |  |  |
| --- | --- | --- | --- |
|  | Seroconverted<br>(n= 103) | Did not seroconvert<br>(n=194) | Not assessed for<br>seroconversion<br>(n=85) |
| Child sex (female) | 53 (51.4) | 97 (50.0) | 40 (47.6) |
| Age of child at HMO<sup>b</sup> measurement (month) | 1.3 (1.2, 1.5) | 1.3 (1.2, 1.4) | 1.3 (1.2, 1.5) |
| Age of child at 1<sup>st</sup> ORV<sup>c</sup> dose (month) | 2.0 (2.0, 2.1) | 2.0 (2.0, 2.1) | 2.0 (2.0, 2.1) |
| Missing date of 1<sup>st</sup> dose | 1 | 2 | 8 |
| Age of child at 2<sup>nd</sup> ORV dose (month) | 4.0 (4.0, 4.2) | 4.0 (4.0, 4.1) | 4.0 (4.0, 4.1) |
| Missing recorded date of 2<sup>nd</sup> dose | 29 | 49 | 20 |
| Maternal secretor phenotype (secretor) | 96 (93.2) | 179 (92.3) | 73 (86.9) |
| Child secretor phenotype (secretor) | 93 (90.3) | 168 (86.6) | 79 (94.0) |
| Child Lewis phenotype |  |  |  |
| Lewis A phenotype | 7 (6.8) | 20 (10.3) | 4 (4.8) |
| Lewis B phenotype | 78 (75.7) | 148 (76.3) | 70 (83.3) |
| Lewis negative | 18 (17.4) | 26 (13.4) | 10 (11.9) |
| Number of older siblings |  |  |  |
| 0 | 51 (49.5) | 91 (46.9) | 37 (44.1) |
| 1 | 38 (36.9) | 71 (36.6) | 30 (35.7) |
| 2 | 10 (9.7) | 22 (11.3) | 9 (10.7) |
| 3 or more | 4 (3.9) | 10 (5.2) | 8 (9.5) |
| Number of household children <3 years old |  |  |  |
| 0 | 2 (1.9) | 10 (5.2) | 5 (6.0) |
| 1 | 59 (57.3) | 114 (58.8) | 44 (52.4) |
| 2 | 42 (40.8) | 70 (36.1) | 35 (41.7) |
| Mother is primary caregiver | 57 (55.3) | 116 (59.8) | 41 (48.8) |
| Home floor construction |  |  |  |
| Dirt | 30 (29.1) | 59 (30.4) | 27 (32.1) |
| Poured cement | 38 (36.9) | 52 (26.8) | 22 (26.2) |
| Cement block or brick | 17 (16.5) | 32 (16.5) | 21 (25.0) |
| Ceramic tiles | 18 (17.5) | 51 (26.3) | 14 (16.7) |
| Mother's age at birth of study child (years) | 23.7 (20.3, 28.9) | 24.0 (19.8, 27.3) | 23.8 (19.9, 27.0) |
| Mother's formal education (years) | 13.0 (10.0, 14.0) | 13.0 (10.0, 17.0) | 13.0 (11.0, 16.0) |
| Duration of any breastfeeding (month) | 22.8 (10.8, 32.1) | 17.5 (8.8, 30.6) | 12.5 (4.8, 24.8) |

<sup>a</sup>IQR, interquartile range<sup>b</sup>HMO, human milk oligosaccharides<sup>c</sup>ORV, oral rotavirus Vaccine

### Rotavirus AGE cumulative incidence

We detected wild-type rotavirus in stools from 69 children experiencing AGE between 1-month post-vaccination and 36 months. We could not type stools with Ct values ≥ 29 (57.3% of infections); genotypes identified included equine-like G3P[8], G8P[8], G8P[nt], G12P[8], and G2P[nt]. The IPT-weighted 36-month rotavirus AGE cumulative incidence was lower in seroconverters (14.3%, 95% CI 7.2, 21.5) than non-seroconverters (19.6%, 95% CI 13.7, 25.4) (Fig 2). Seroconverters were also approximately 5 months older (approximately 12 months of age) at their first rotavirus AGE compared to non-seroconverters. Children with positive anti-rotavirus IgA titers before vaccination also tended to have a higher IPT-weighted cumulative incidence of rotavirus AGE (17.2%, 95% CI 6.0, 28.4) than seroconverters.

**Fig 2.**
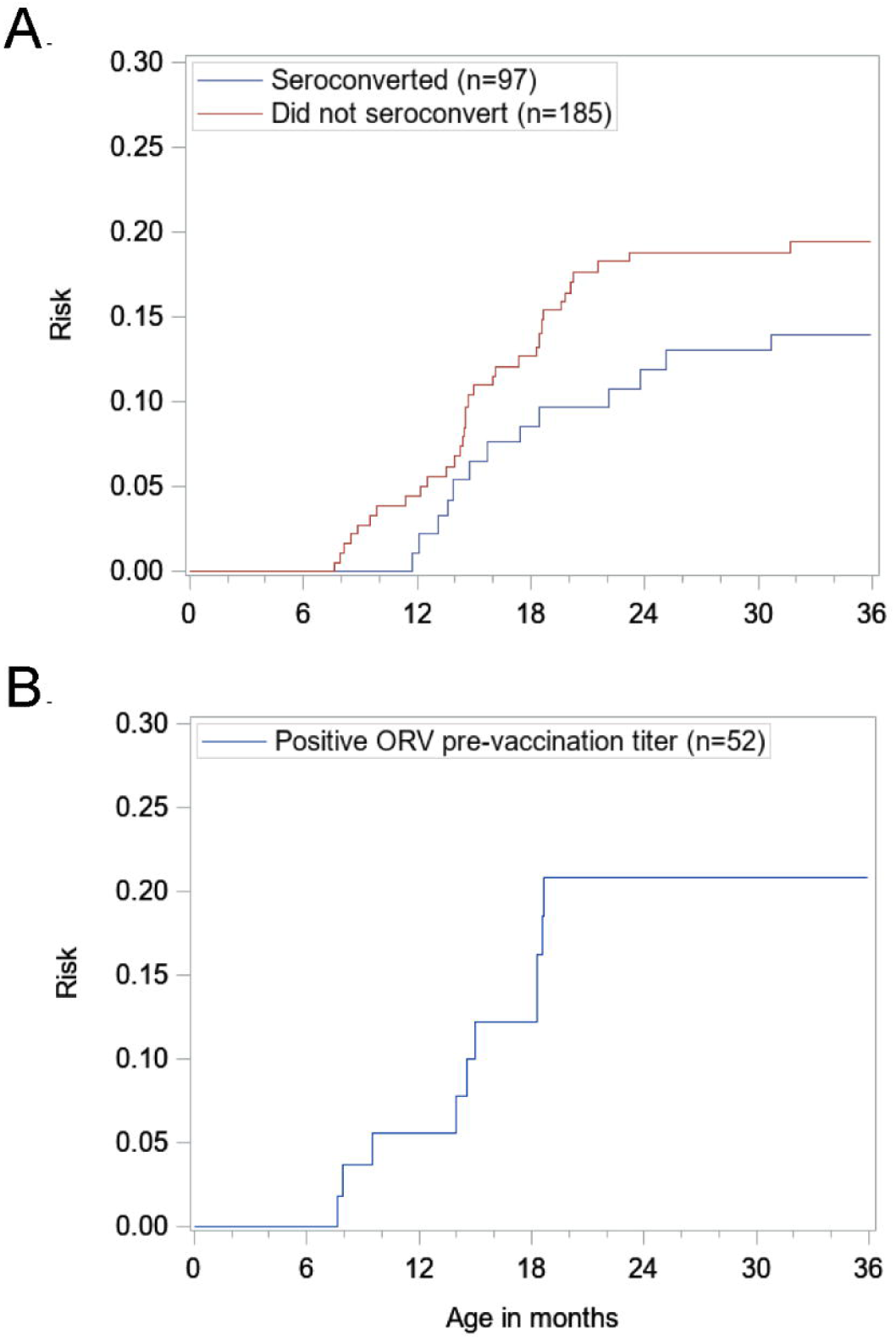
Cumulative incidence (risk) of rotavirus acute gastroenteritis (AGE) and time-to-first rotavirus AGE for a) stratified by seroconversion status among those assessed for seroconversion (n=282) and b) children with positive pre-vaccination titers to Rotarix GlP[S] antigen (n=52). **Footnote:** Follow-up initiated the day of post-vaccination titer quantification in serum (−1 month after the 2nd oral rotavirus vaccine dose). Cumulative incidence was detennined using a Kaplan-Meier survival function assuming no competing risks. Cumulative incidence curves for children who did and *did* not seroconvert were weighted for the following hypothesized confounders of the relationship between seroconversion and rotavirusAGE: weeks of exclusive breastfeeding prior to seroconversion ascertainment, currently breastfeeding at time of seroconversion ascertainment, mother’s secretor phenotype, child’s secretor and Lewis phenotypes. The cumulative incidence for participants seropositive at baseline was weighted for the following hypothesized confounders: mother’s secretor phenotype, child’s secretor and Lewis phenotypes. Nine non-seroconverters and 6 seroconverters were not included because the participants’ censoring dates occurred before the date in which their serum was collected for seroconversion status.

### Associations between HMO concentrations and seroconversion probability

Among all cohort children who were assessed by serology, receiving maternal milk with higher concentrations of 2’FL was positively associated with seroconversion, although with confidence intervals crossing the null, while higher concentrations of the HMOs 3’siallylactose (3’SL), lacto-N-tetraose (LNT), lacto-N-fucopentaose (LNFP-I), LNFP-II, sialyllacto-N-tetraose c (LSTc), difucosyllacto-N-tetraose (DFLNT) and difucosyllactose (DFLNH) were negatively associated with seroconversion (Fig 3). For example, children receiving the middle concentration of 2’FL were 31% (95% CI, −10%, 92%) more likely to seroconvert to RV1, as compared to children receiving the lowest concentration of 2’FL.

**Fig 3.**
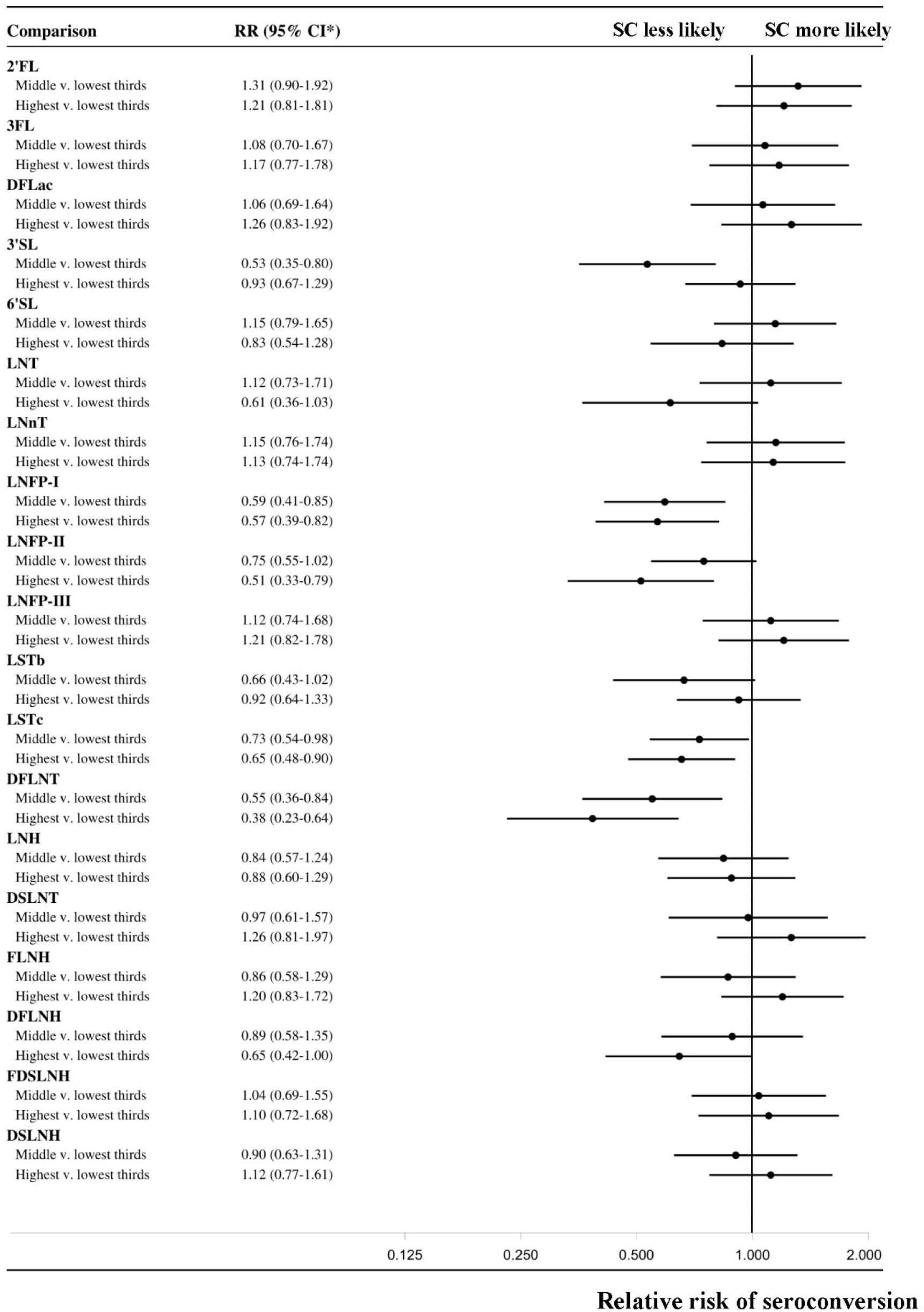
Association between human milk oligosaccharide (HMO) concentrations and seroconversion (SC) to the monovalent rotavirus vaccine (RVl), adjusted analysis of all. **footnote:** Adjusted 5-month risk ratio (RR) and 95% confidence interval (CI) of RV! seroconversion by maternal HMO concentration among 297 children for whom seroconversion was determined. Estimates of RR were weighted for the inverse probability of treatment (IPTW) to account for bias from confounding. RR estimates are represented by black circles and width of 95% confidence intervals (Cls) by solid black lines. Null RR is 1.00, marked with a solid line. RR<l indicates that participants consuming the intem1ediate or highest concentration of a given I-IMO were less likely to seroconvert compared to participants consuming the lowest concentrations of a given HMO. RR>1 indicates the opposite relationship between I-IMO concentration and the probability of seroconversion.

In the stratified analysis of secretor children, receiving higher concentrations of the HMOs 2’FL, 3-fucosyllactose (3FL), DFLac, and disialyllacto-N-hexaose (DSLNH) were positively associated with seroconversion (Fig 4), while higher concentrations of 3’SL, LNFP-I, sialyllacto-N-tetraose b (LSTb), LSTc, and DFLNT were negatively associated with seroconversion (Fig 4). We did not apply IPCWs as all children were still breastfeeding at the post-vaccination IgA time-point (5 months). We could not assess outcomes within the non-secretor subgroup due to the small subgroup size.

**Fig 4.**
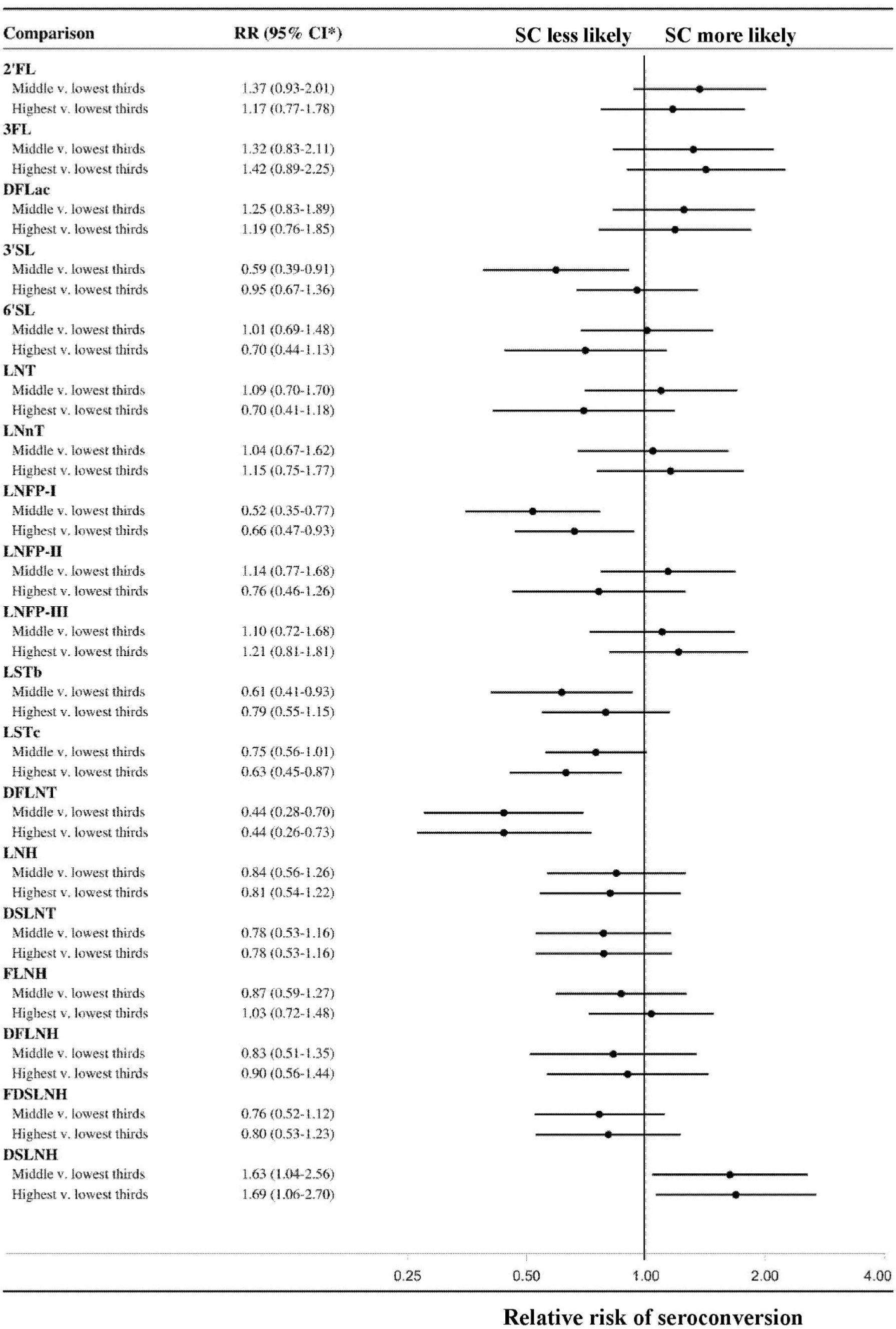
Association between human milk oligosaccharide (HMO) concentration and seroconv ersion (SC) to the monovalent rotavirus vaccine (RVl), adjusted analysis among. **footnote:** Adjusted 5-month risk ratio (RR) and 95% confidence interval (CI) of RV] seroconversion by maternal HMO concentration among 246 secretor children for whom seroconversion was detennined.

### Associations between HMO concentrations and risk of rotavirus AGE

Among all children, receiving higher concentrations of LNFP-I was strongly associated with increased rotavirus AGE risk and higher concentrations of DFLNH was associated with decreased risk, although with confidence intervals crossing the null (Fig 5). For example, children receiving the highest concentration of DFLNH had a difference in the absolute risk of rotavirus AGE that was 15.8 percentage points lower (95% CI, −5.4%-37.0%) than children receiving the lowest concentration of DFLNH. In the stratified models including only secretor children, receiving higher concentrations of 6’ sialyllactose (6’SL), LNT, LNFP-I, and DSLNT were associated with increased rotavirus AGE risk and higher concentrations of DFLNH was associated with decreased risk (Fig 6). These analyses of HMO concentrations and rotavirus AGE incidence were similar to the analyses in which children were censored upon weaning (Supplemental Figs S10 and S11).

**Fig 5.**
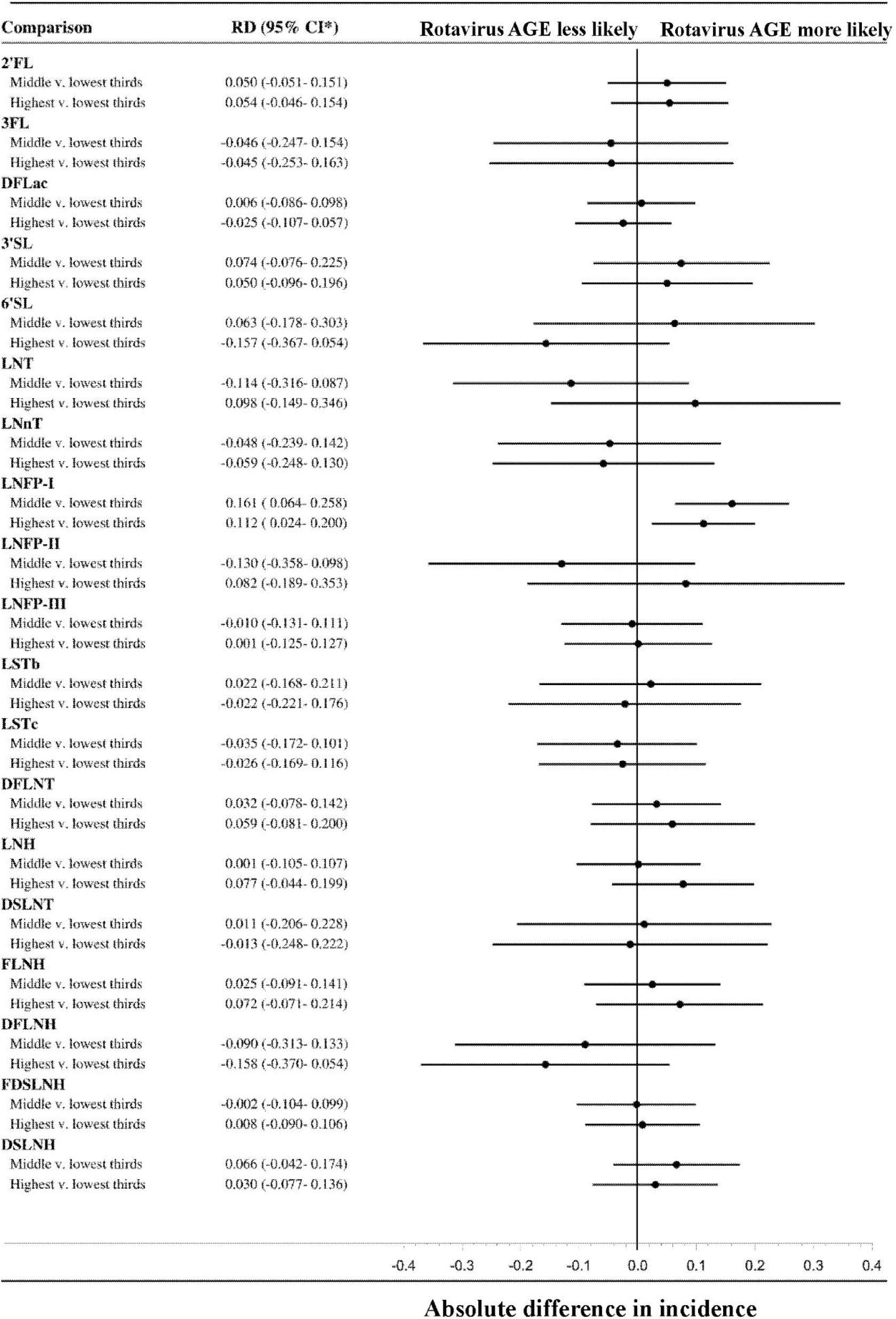
Association between human milk oligosaccharide (HMO) concentration and rotavirus acute gastroenteritis (AGE) in the first 36 months of life, adjusted analysis of. **footnote:** Adjusted 36-month risk difference (RD) and 95% CI of rotavirus AGE by maternal HMO concentration among all 282 cohort children with maternal milk HMOs characterized and who had remained in the study c:1 month after vaccination and had been characterized for seroconversion. Estimates were weighted with the inverse probability of treatment weights (IPTWs) to account for potential bias from confounding. Null RD is 0.00, marked with a solid line. RD<0 indicates that participants consuming the intennediate or highest concentration of a given HMO were less likely to experience rotavirusAGE compared to participants consuming the lowest concentrations of a given HMO. RD>0 indicates the opposite relationship between HMO concentration and rotavims AGE risk.

**Fig 6.**
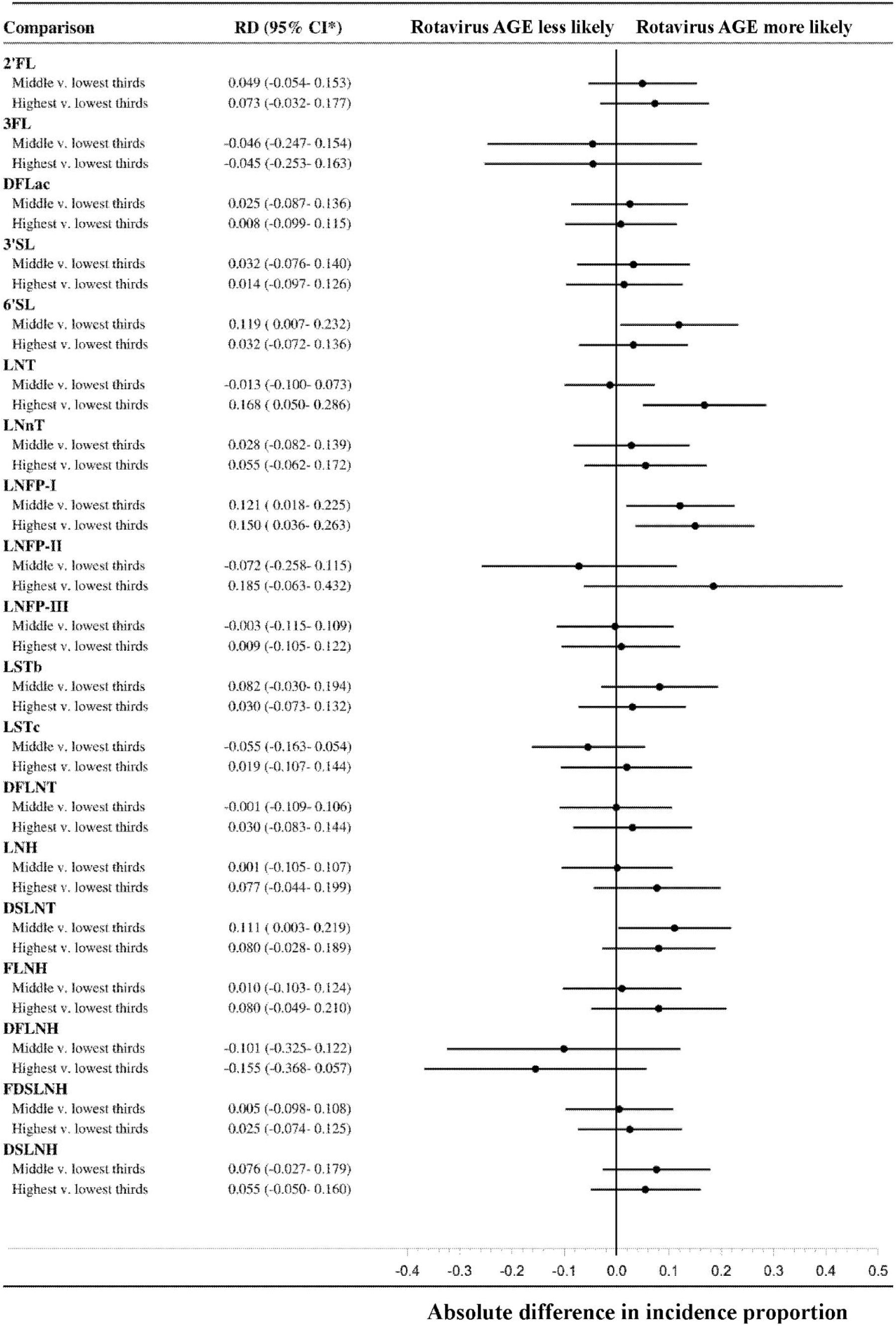
Association between human milk oligosaccharide (HMO) concentration and rotavirus acute gastroenteritis (AGE), adjusted analysis of secretor children. **footnote:** Adjusted 36-month risk difference (RD) and 95% CI ofrotavirnsAGE by maternal HMO composition among 246 secretor cohort children with HMOs characterized who had remained in the study 2: I-month post-vaccination and for whom seroconversion was detem1ined.

## Discussion

In this population-based cohort in Central America, receiving higher concentrations of several HMOs, most notably, LNFP-I, LNFP-II, LSTc, and DFLNT among all children, were negatively associated with RV1 seroconversion (Fig 3). While the biological mechanisms behind this finding are not entirely clear from this epidemiological study, it is possible that these HMOs may have reduced vaccine seroconversion by acting as soluble attachment factors^8,9^, or by promoting gut commensals that could contribute to decreased vaccine virus replication. Conversely, higher concentrations of the fucosylated HMO 2’FL was positively associated with RV1 seroconversion, although with confidence intervals crossing the null. Our results were somewhat unexpected, given the evidence from in vitro models showing that 2’FL decreases G1P[8] infectivity^34^, suggesting that higher concentrations of 2’FL would decrease RV1 seroconversion. In addition, prior studies from Bangladesh and Zimbabwe identified lower RV1 seroconversion in children of secretor mothers^8,9^, who would be expected to produce higher concentrations of 2’FL in their milk than non-secretor mothers^7,26^. This suggests that factors other than high 2’FL concentrations in secretor mothers may be responsible for these prior findings.

Children receiving milk with higher concentrations of the fucosylated HMO, DFLNH, had a lower risk of rotavirus AGE over the first 3 years of life. In contrast, receiving higher concentrations of two other fucosylated HMOs, LNFP-1 and DFLNT, were associated with both a lower likelihood of RV1 seroconversion, and a correspondingly higher risk of rotavirus AGE. While fucosylated and sialylated HMOs were more likely to be associated with the outcomes examined as compared to neutral non-fucosylated HMOs, we did not find a “class effect” of HMOs on rotavirus susceptibility, i.e. HMOs within the same structural category had differing associations. Also, not all HMOs that were associated with RV1 seroconversion were also associated with rotavirus AGE risk over the first 3 years of life. For example, while higher 2’FL was associated with increased seroconversion, it was not associated with a lower risk of rotavirus AGE. It is possible that the longer-term effects of HMOs on rotavirus AGE risk may be confounded by many other exposures that occur after the protection from RV1 wanes around the child’s first birthday. It is also possible that 2’FL promotes gut bacteria and enteric immune cells that enhance the infectivity of both vaccine-strain Rotarix G1P[8] (promoting seroconversion) and wild-type rotavirus, minimizing the beneficial effect of increased seroconversion^12,35,36^.

We did not find notable differences in our results for rotavirus AGE risk when children were censored after weaning. This suggests that the effects of HMOs on rotavirus AGE are not limited to their immediate action as decoy receptors, but may have broader mechanisms of action, for example, in shaping gut microbiome composition.

Only 1/3 of the infants in this cohort seroconverted to RV1, comparable to countries with the highest global under-5 mortality^37^. Importantly, RV1 seroconversion, defined as ≥ 4-fold increase in serum IgA titers to vaccine virus, was associated with a delay in the onset of rotavirus AGE by approximately 5 months. After 12 months of age, children experienced a similar rate of rotavirus AGE regardless of seroconversion status (Fig 2). Therefore, RV1 seroconversion functioned as a robust—yet transient—vaccine correlate of protection against rotavirus AGE of any severity in our cohort. Unlike other studies, seroconversion frequency in this cohort did not differ by maternal or child secretor status^8,9,38–41^, though our results may be limited by the low proportion of non-secretors in our population.

We also identified very low RV1 seroconversion in the 14.8% of infants with elevated pre-vaccination IgA titers to Rotarix G1P[8] antigen. A US-based study identified a similar relationship in children with pre-vaccination anti-RV IgG^38^ (in that case, likely representing maternally-acquired immunity). Explanations for positive pre-vaccination IgA titers include infection with wild-type rotavirus prior to vaccination and transmission of vaccine-derived rotavirus from recently vaccinated contacts via vaccine shedding in stool^42–44^.

Our results clearly show that pre-vaccination anti-rotavirus IgA dampens the RV1 response. Nonetheless, even after removing infants with anti-rotavirus IgA prior to vaccination, seroconversion frequency remained low at 42.3%, suggesting multiple mechanisms contributed to low seroconversion.

A limitation of this study is that we did not directly measure anti-rotavirus antibodies in mothers’ milk; in addition to having a different distribution of HMOs, secretor mothers’ milk may have higher levels of anti-rotavirus antibodies, as secretors have increased susceptibility to rotavirus. To account for this, we adjusted for maternal secretor status in our models. Interestingly, some of the beneficial associations between HMOs and the outcomes we examined were found for HMOs that have higher levels in the milk of non-secretor mothers as compared to secretor mothers, such as 3FL and DFLNH^26^. Another limitation, common to many studies of the health benefits of human milk, is that we did not measure the daily “dose” of human milk received by the children, in terms volume of human milk consumed. While consumed milk volume can be estimated by obtaining precise weights of the child before and after each feeding, this is not logistically feasible in a large cohort and adds a large time burden for participating mothers. Finally, unmeasured child diet or environmental variables could have confounded associations between seroconversion, pre-vaccination IgA, and rotavirus AGE incidence, and unmeasured maternal diet could have confounded models of HMOs, seroconversion, and rotavirus AGE^45^.

Our study is strengthened by its population-based prospective cohort design, which includes weekly household surveillance over three years in a Central American community with a high AGE burden. While others have assessed maternal and child secretor statuses and ORV seroconversion, or maternal and child secretor statuses and rotavirus AGE, our study investigates both relationships jointly, while incorporating the role of HMOs on both outcomes in breastfeeding and weaned children.

Our findings suggest that 2’FL could promote vaccine effectiveness, though additional studies are needed to confirm its mechanism of protection. As higher concentrations of 2’FL was positively associated with seroconversion, it represents a viable strategy to improve rotavirus vaccine effectiveness as a supplement for breastfeed infants, or for inclusion in infant formulas^46^, in settings where rotavirus burden is highest.

Nonetheless, our findings suggest that having higher concentrations of several HMOs may contribute to poorer vaccine response, despite the well-established health benefits of breastfeeding. Additionally, no HMOs were associated with both seroconversion *and* protection against gastroenteritis, suggesting that both ORV seroconversion and rotavirus gastroenteritis outcomes must be monitored in efficacy trials of HMOs and rotavirus prevention. While numerous trials have found that HMO-supplemented formula promotes healthy infant growth and biomarkers mimicking breastfed infants^13^, our data suggest that HMOs are not risk-free, and that more clinical evidence of their benefit is needed before they are added to infant formulas in regions with poor ORV efficacy and high rotavirus disease burden.

In the future, mixed vaccine schedules including both oral and parental vaccines^47,48^ may be one option of increasing immunogenicity while avoiding interference from human milk components. Our findings of the short-lived protection conferred by RV1 highlights the need to improve the duration of protection of ORVs, with approaches such as booster doses^49^ or new vaccines that promote long-lived protection. This is particularly important because even children who seroconverted to the vaccine experienced high rates of rotavirus AGE in the second year of life. Without new interventions, rotavirus will likely remain the leading cause of worldwide diarrhea-related mortality in young children.

## Data Availability

All data produced in the present study are available upon reasonable request to the authors.

## Acknowledgments

We especially thank the participants, family members, and laboratory and field teams who made the SAGE cohort possible, especially those involved in the fieldwork and lab-work components of the study: Merling Balmaceda, Vanessa Bolaños, Nancy Corea, Jhosselyng Delgado, Marvel Fuentes, Yadira Hernandez, Llurvin Madriz, Patricia Mendez, Yuvielka Martinez, Maria Mendoza, Ruth Neira, Xiomara Obando, Veronica Pravia, Yorling Picado, Aura Scott, and Mileydis Soto.

## Disclaimer

The findings and conclusions in this report are those of the authors and do not necessarily represent the views of the US Centers for Disease Control and Prevention.

## Conflict of Interest Disclosures (includes financial disclosures)

LB is a co-inventor on patent applications related to the use of HMOs in preventing NEC and other inflammatory diseases. The other authors have no conflicts of interest to disclose.

## Role of Funder/Sponsor (if any)

All funding sources had no role in the design and conduct of the study, nor any restrictions regarding publication.

## Abbreviations

HMO: human milk oligosaccharide
ORV: oral rotavirus vaccines
RV: rotavirus
LMICs: low-and-middle-income countries
IgA: immunoglobulin A
AGE: acute gastroenteritis episode
HICs: high-income countries
HBGA: histo-blood group antigen
FUT2: fucosyltransferase-2
FUT3: fucosyltransferase-3
SAGE: Sapovirus Acute GastroEnteritis
UNAN: Universidad Nacional Autónoma de Nicaragua
qPCR: quantitative polymerase chain reaction
ELISA: enzyme-linked immunosorbent assay
HPLC/UPLC-FL: high-and-ultra-high pressure liquid chromatography after fluorescent labeling
IQR: interquartile range
IPTW: inverse probability of treatment weighting
RR: relative risks
RDs: risk differences
CIs: confidence intervals
DAG: directed acyclic graph
IPCWs: inverse probability of censoring weights
2’FL: 2’fucosyllactose
3FL: 3-fucosyllactose
DFLac: difucosyllactose
3’SL: 3’siallylactose
6’SL: 6’ sialyllactose
LNT: lacto-N-tetraose
LNnT: lacto-N-neotetraose
LNFP-I: fucosylated lacto-N-fucopentaose-I
LNFP-II: fucosylated lacto-N-fucopentaose-II
LNFP-III: fucosylated lacto-N-fucopentaose-III
LSTb: sialyllacto-N-tetraose b
LSTc: sialyllacto-N-tetraose c
DFLNT: difucosyllacto-N-tetraose
LNH: lacto-N-hexaose
DSLNT: disialyllacto-N-tetraose
FLNH: fucosyllacto-N-hexaose
DFLNH: difucosyllacto-N-hexaose
FDSLNH: fucosyl-disialyllacto-N-hexose
DSLNH: disialyllacto-N-hexaose

