## Supplementary Information for "Human milk oligosaccharides, oral rotavirus vaccine seroconversion, and rotavirus gastroenteritis risk in a vaccinated birth cohort"

**Supplement:**

**Description of IPTW calculation.**

IPTWs were generated for seroconversion/rotavirus AGE, seropositivity/rotavirus AGE, HMO/seroconversion and HMO/rotavirus AGE associations. We calculated IPTWs by dividing the probability of seroconversion or HMO exposure status (highest, middle, lowest concentrations) in an intercept-only logistic regression by the probability of seroconversion or HMO exposure status (highest, middle, lowest concentrations) conditional on potential confounders described in the text. Numerators and denominators were calculated with logistic regressions with a multinomial link function to calculate weights for all three exposure categories (low, middle, high) in one model. Then, we included weighted participants in 2 comparisons (middle v. low and high v. low).

Confounders were identified via subject matter expertise and a review of the literature, and a minimally sufficient adjustment set was identified using a causal DAG. In addition to the confounders listed below, we also adjusted for HMOs that correlated with the HMO exposure containing a correlation coefficient between 0.5-1 by adding them to the IPTW denominator. The functional forms of the numerator and denominator models for the IPTWs are as follows:

**Association between seroconversion and rotavirus AGE:**

**Numerator:**

$\text{logit}(E(y)) = \beta_0$ , where y = seroconversion

**Denominator:**

$\text{logit}(E(y)) = \beta_0 + \beta_1 X_1 + \beta_2 X_2 + \beta_3 X_3 + \beta_4 X_4 + \beta_5 X_5 + \beta_6 X_{6\ldots} \beta_n X_n$

where $X_1$ = weeks of exclusive breastfeeding prior to seroconversion determination

$X_2$ = any breastfeeding at time of seroconversion

$X_3$ = mother's secretor phenotype (milk)

$X_4$ = child's secretor phenotype (saliva)

$X_5$ = child's Lewis phenotypes (saliva)

$X_{6\ldots} X_n$ = HMOs correlated with primary HMO exposure with Pearson correlation coefficient 0.5-1.

**Association between seropositivity at baseline and rotavirus AGE:**

**Numerator:**

$\text{logit}(E(y)) = \beta_0$ , where y = seropositive at baseline

**Denominator:**

$\text{logit}(E(y)) = \beta_0 + \beta_1 X_1 + \beta_2 X_2 + \beta_3 X_3$

where y=seropositive at baseline

$X_1$ = mother's secretor phenotype (milk)

$X_2$ = child's secretor phenotype (saliva)

$X_3$ = child's Lewis phenotypes (saliva)

**Association between HMO concentration and seroconversion and HMO concentration and rotavirus AGE:**

**Numerators:**

<u>Comparison of middle v. lowest HMO concentrations:</u>

$\text{logit}(E(y)) = \beta_0$ , where y = middle HMO concentration

<u>Comparison of highest v. lowest HMO concentrations:</u>

$\text{logit}(E(y)) = \beta_0$ , where y = highest HMO concentration

**Denominators:**

<u>Comparison of middle v. lowest HMO concentrations:</u>

$\text{logit}(E(y)) = \beta_0 + \beta_1 X_1 + \beta_2 X_2 + \beta_3 X_3 + \beta_4 X_4 + \beta_5 X_5 + \beta_6 X_6$ ,

where y=middle HMO concentration

$X_1$ = cumulative weeks of any breastfeeding at HMO measurement

$X_2$ = child's age (approximating lactation stage) at HMO measurement

$X_3$ = maternal age at the child's birth

$X_4$ = number of child's older siblings (approximating parity)

$X_5$ = child secretor phenotype

$X_6$ = child Lewis phenotype

<u>Comparison of highest v. lowest HMO concentrations:</u>

$\text{logit}(E(y)) = \beta_0 + \beta_1 X_1 + \beta_2 X_2 + \beta_3 X_3 + \beta_4 X_4 + \beta_5 X_5 + \beta_6 X_6$ ,

where y=highest HMO concentration

$X_1$ = cumulative weeks of any breastfeeding at HMO measurement

$X_2$ = child's age (approximating lactation stage) at HMO measurement

$X_3$ = maternal age at the child's birth

$X_4$ = number of child's older siblings (approximating parity)

$X_5$ = child secretor phenotype

$X_6$ = child Lewis phenotype

**Description of IPCW calculation.**

IPCWs were generated for the HMO/rotavirus AGE associations. We calculated IPCWs by dividing the probability of weaning on uncensored participants in an intercept-only logistic regression by the probability of weaning conditional on potential confounders described in the text. Logistic regressions were fit on uncensored participants. As weaning is a time-varying condition, the values of time-varying confounders and the predicted probabilities of weaning for each participant were updated with each weekly visit. Confounders were identified via subject matter expertise and a review of the literature, and a minimally sufficient adjustment set was identified using a causal DAG. The functional forms of the numerator and denominator models for the IPCWs are as follows:

**Numerator:**

$\text{logit}(E(y)) = \beta_0$ , where y = weaned within the last 2 weeks

**Denominator:**

$\text{logit}(E(y)) = \beta_0 + t + X_1 + X_1 t + X_2 + X_2 t + X_3 + X_3 t + X_4 + X_4 t + X_5 + X_6 + X_7 + X_8$

where $t$ = weeks since study entry

$X_1$ = siblings <3 years of age in home (yes/no)

$X_2$ = primary caregiver to child is mother v. other household member

$X_3$ = vomiting or diarrhea in past 3 days

$X_4$ = household water supply interrupted in current month

$X_5$ = household floor type (approximating household poverty)

$X_6$ = number of siblings in home

$X_7$= maternal age at child's birth

$X_8$= years of maternal formal education

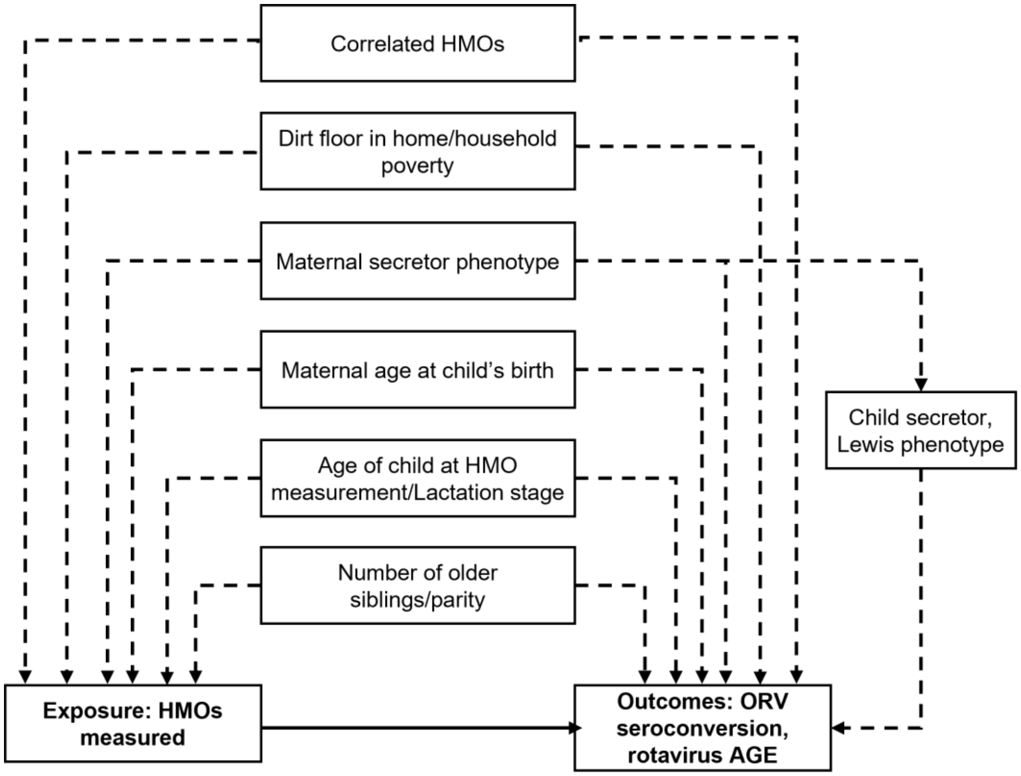


**Supplementary Figure S1:** Simplified directed acyclic graph (DAG) identifying the minimally sufficient adjustment set of observed confounders of the causal effect of composition of HMOs in milk consumed in early life on the outcomes of ORV seroconversion and 36-month rotavirus AGE incidence, represented by a solid black arrow. Using the software daggity.net[1], we identified this adjustment set of confounders to include parity, age of child at HMO measurement (as a proxy for lactation stage), maternal age at study child's birth, maternal secretor phenotype, presence of dirt floors in the home as a proxy for household poverty[2], and correlated HMOs with Pearson r 0.5-1 as described previously. Child HBGA phenotype was identified as an effect measure modifier but was also included in IPTW calculations.

| | 2'FL | 3FL | DFLac | 3'SL | 6'SL | LNT | LNnT | LNFP-I | LNFP-II | LNFP-III | LSTb | LSTc | DFLNT | LNH | DSLNT | FLNH | DFLNH | FDSLNH | DSLNH |
|---|---|---|---|---|---|---|---|---|---|---|---|---|---|---|---|---|---|---|---|
| 2'FL | | -0.34566 | 0.18581 | 0.17315 | 0.03145 | -0.31345 | 0.21231 | 0.54489 | -0.38522 | 0.01089 | -0.13325 | 0.28785 | 0.00981 | 0.07653 | -0.01759 | -0.04966 | -0.35051 | -0.3015 | 0.2479 |
| | 2'FL | 3FL | DFLac | 3'SL | 6'SL | LNT | LNnT | LNFP-I | LNFP-II | LNFP-III | LSTb | LSTc | DFLNT | LNH | DSLNT | FLNH | DFLNH | FDSLNH | DSLNH |
| 3FL | -0.34566 | | 0.04168 | 0.12304 | 0.1826 | 0.21553 | -0.01125 | -0.42792 | 0.75892 | -0.18368 | 0.09323 | -0.10609 | 0.18971 | 0.12402 | 0.03543 | 0.21588 | 0.69348 | 0.7528 | 0.16008 |
| | 2'FL | 3FL | DFLac | 3'SL | 6'SL | LNT | LNnT | LNFP-I | LNFP-II | LNFP-III | LSTb | LSTc | DFLNT | LNH | DSLNT | FLNH | DFLNH | FDSLNH | DSLNH |
| DFLac | 0.18581 | 0.04168 | | 0.6038 | 0.09865 | -0.12989 | 0.11731 | 0.09245 | 0.04682 | 0.08929 | 0.17855 | 0.33851 | 0.69546 | -0.08768 | 0.35625 | -0.3753 | 0.02374 | -0.17744 | 0.20426 |
| | 2'FL | 3FL | DFLac | 3'SL | 6'SL | LNT | LNnT | LNFP-I | LNFP-II | LNFP-III | LSTb | LSTc | DFLNT | LNH | DSLNT | FLNH | DFLNH | FDSLNH | DSLNH |
| 3'SL | 0.17315 | 0.12304 | 0.6038 | | 0.23859 | 0.04816 | 0.24096 | 0.13558 | 0.20891 | -0.04455 | 0.21727 | 0.34299 | 0.56683 | 0.02365 | 0.41869 | -0.15258 | 0.18189 | -0.02541 | 0.34225 |
| | 2'FL | 3FL | DFLac | 3'SL | 6'SL | LNT | LNnT | LNFP-I | LNFP-II | LNFP-III | LSTb | LSTc | DFLNT | LNH | DSLNT | FLNH | DFLNH | FDSLNH | DSLNH |
| 6'SL | 0.03145 | 0.1826 | 0.09865 | 0.23859 | | 0.5801 | 0.26811 | 0.37179 | 0.35768 | 0.15895 | 0.41245 | 0.58675 | 0.30857 | 0.27096 | 0.53409 | 0.37809 | 0.2999 | 0.17189 | 0.64489 |
| | 2'FL | 3FL | DFLac | 3'SL | 6'SL | LNT | LNnT | LNFP-I | LNFP-II | LNFP-III | LSTb | LSTc | DFLNT | LNH | DSLNT | FLNH | DFLNH | FDSLNH | DSLNH |
| LNT | -0.31345 | 0.21553 | -0.12989 | 0.04816 | 0.5801 | | 0.41461 | 0.22186 | 0.55958 | 0.11483 | 0.50818 | 0.20288 | 0.16829 | 0.33104 | 0.55047 | 0.47787 | 0.4543 | 0.27791 | 0.22578 |
| | 2'FL | 3FL | DFLac | 3'SL | 6'SL | LNT | LNnT | LNFP-I | LNFP-II | LNFP-III | LSTb | LSTc | DFLNT | LNH | DSLNT | FLNH | DFLNH | FDSLNH | DSLNH |
| LNnT | 0.21231 | -0.01125 | 0.11731 | 0.24096 | 0.26811 | 0.41461 | | 0.27704 | 0.2586 | 0.06259 | 0.26788 | 0.46555 | 0.34632 | 0.53716 | 0.39176 | 0.2389 | 0.23924 | 0.0513 | 0.21925 |
| | 2'FL | 3FL | DFLac | 3'SL | 6'SL | LNT | LNnT | LNFP-I | LNFP-II | LNFP-III | LSTb | LSTc | DFLNT | LNH | DSLNT | FLNH | DFLNH | FDSLNH | DSLNH |
| LNFP-I | 0.54489 | -0.42792 | 0.09245 | 0.13558 | 0.37179 | 0.22186 | 0.27704 | | -0.25214 | 0.30895 | 0.27532 | 0.44833 | 0.06062 | 0.12844 | 0.41439 | 0.04371 | -0.24456 | -0.36911 | 0.26885 |
| | 2'FL | 3FL | DFLac | 3'SL | 6'SL | LNT | LNnT | LNFP-I | LNFP-II | LNFP-III | LSTb | LSTc | DFLNT | LNH | DSLNT | FLNH | DFLNH | FDSLNH | DSLNH |
| LNFP-II | -0.38522 | 0.75892 | 0.04682 | 0.20891 | 0.35768 | 0.55958 | 0.2586 | -0.25214 | | -0.00685 | 0.40915 | 0.13766 | 0.35451 | 0.22259 | 0.37155 | 0.28334 | 0.88086 | 0.68653 | 0.29092 |
| | 2'FL | 3FL | DFLac | 3'SL | 6'SL | LNT | LNnT | LNFP-I | LNFP-II | LNFP-III | LSTb | LSTc | DFLNT | LNH | DSLNT | FLNH | DFLNH | FDSLNH | DSLNH |
| LNFP-III | 0.01089 | -0.18368 | 0.08929 | -0.04455 | 0.15895 | 0.11483 | 0.06259 | 0.30895 | -0.00685 | | 0.54468 | 0.38951 | 0.13148 | 0.17654 | 0.26081 | 0.06619 | -0.07311 | -0.11381 | 0.11999 |
| | 2'FL | 3FL | DFLac | 3'SL | 6'SL | LNT | LNnT | LNFP-I | LNFP-II | LNFP-III | LSTb | LSTc | DFLNT | LNH | DSLNT | FLNH | DFLNH | FDSLNH | DSLNH |
| LSTb | -0.13325 | 0.09323 | 0.17855 | 0.21727 | 0.41245 | 0.50818 | 0.26788 | 0.27532 | 0.40915 | 0.54468 | | 0.4235 | 0.44777 | 0.2256 | 0.67946 | 0.14878 | 0.31678 | 0.07493 | 0.29224 |
| | 2'FL | 3FL | DFLac | 3'SL | 6'SL | LNT | LNnT | LNFP-I | LNFP-II | LNFP-III | LSTb | LSTc | DFLNT | LNH | DSLNT | FLNH | DFLNH | FDSLNH | DSLNH |
| LSTc | 0.28785 | -0.10609 | 0.33851 | 0.34299 | 0.58675 | 0.20288 | 0.46555 | 0.44833 | 0.13766 | 0.38951 | 0.4235 | | 0.51357 | 0.37797 | 0.5081 | 0.04722 | 0.10012 | -0.09393 | 0.57163 |
| | 2'FL | 3FL | DFLac | 3'SL | 6'SL | LNT | LNnT | LNFP-I | LNFP-II | LNFP-III | LSTb | LSTc | DFLNT | LNH | DSLNT | FLNH | DFLNH | FDSLNH | DSLNH |
| DFLNT | 0.00981 | 0.18971 | 0.69546 | 0.56683 | 0.30857 | 0.16829 | 0.34632 | 0.06062 | 0.35451 | 0.13148 | 0.44777 | 0.51357 | | 0.15181 | 0.52764 | -0.19997 | 0.27655 | 0.0002 | 0.26065 |
| | 2'FL | 3FL | DFLac | 3'SL | 6'SL | LNT | LNnT | LNFP-I | LNFP-II | LNFP-III | LSTb | LSTc | DFLNT | LNH | DSLNT | FLNH | DFLNH | FDSLNH | DSLNH |
| LNH | 0.07653 | 0.12402 | -0.08768 | 0.02365 | 0.27096 | 0.33104 | 0.53716 | 0.12844 | 0.22259 | 0.17654 | 0.2256 | 0.37797 | 0.15181 | | 0.23998 | 0.61331 | 0.23429 | 0.34292 | 0.2602 |
| | 2'FL | 3FL | DFLac | 3'SL | 6'SL | LNT | LNnT | LNFP-I | LNFP-II | LNFP-III | LSTb | LSTc | DFLNT | LNH | DSLNT | FLNH | DFLNH | FDSLNH | DSLNH |
| DSLNT | -0.01759 | 0.03543 | 0.35625 | 0.41869 | 0.53409 | 0.55047 | 0.39176 | 0.41439 | 0.37155 | 0.26081 | 0.67946 | 0.5081 | 0.52764 | 0.23998 | | 0.15812 | 0.33089 | -0.00178 | 0.4205 |
| | 2'FL | 3FL | DFLac | 3'SL | 6'SL | LNT | LNnT | LNFP-I | LNFP-II | LNFP-III | LSTb | LSTc | DFLNT | LNH | DSLNT | FLNH | DFLNH | FDSLNH | DSLNH |
| FLNH | -0.04966 | 0.21588 | -0.3753 | -0.15258 | 0.37809 | 0.47787 | 0.2389 | 0.04371 | 0.28334 | 0.06619 | 0.14878 | 0.04722 | -0.19997 | 0.61331 | 0.15812 | | 0.30609 | 0.54599 | 0.32553 |
| | 2'FL | 3FL | DFLac | 3'SL | 6'SL | LNT | LNnT | LNFP-I | LNFP-II | LNFP-III | LSTb | LSTc | DFLNT | LNH | DSLNT | FLNH | DFLNH | FDSLNH | DSLNH |
| DFLNH | -0.35051 | 0.69348 | 0.02374 | 0.18189 | 0.2999 | 0.4543 | 0.23924 | -0.24456 | 0.88086 | -0.07311 | 0.31678 | 0.10012 | 0.27655 | 0.23429 | 0.33089 | 0.30609 | | 0.72694 | 0.33712 |
| | 2'FL | 3FL | DFLac | 3'SL | 6'SL | LNT | LNnT | LNFP-I | LNFP-II | LNFP-III | LSTb | LSTc | DFLNT | LNH | DSLNT | FLNH | DFLNH | FDSLNH | DSLNH |
| FDSLNH | -0.3015 | 0.7528 | -0.17744 | -0.02541 | 0.17189 | 0.27791 | 0.0513 | -0.36911 | 0.68653 | -0.11381 | 0.07493 | -0.09393 | 0.0002 | 0.34292 | -0.00178 | 0.54599 | 0.72694 | | 0.28436 |
| | 2'FL | 3FL | DFLac | 3'SL | 6'SL | LNT | LNnT | LNFP-I | LNFP-II | LNFP-III | LSTb | LSTc | DFLNT | LNH | DSLNT | FLNH | DFLNH | FDSLNH | DSLNH |
| DSLNH | 0.2479 | 0.16008 | 0.20426 | 0.34225 | 0.64489 | 0.22578 | 0.21925 | 0.26885 | 0.29092 | 0.11999 | 0.29224 | 0.57163 | 0.26065 | 0.2602 | 0.4205 | 0.32553 | 0.33712 | 0.28436 | |

**Supplementary Figure S2.** Correlation matrix between all 19 HMOs in all cohort children assessed for seroconversion (n=297). Numeric values represent Pearson correlation coefficients.

| | 2'FL | 3FL | DFLac | 3'SL | 6'SL | LNT | LNnT | LNFP-I | LNFP-II | LNFP-III | LSTb | LSTc | DFLNT | LNH | DSLNT | FLNH | DFLNH | FDSLNH | DSLNH |
|---|---|---|---|---|---|---|---|---|---|---|---|---|---|---|---|---|---|---|---|
| **2'FL** | | -0.31399 | 0.14079 | 0.14173 | 0.01017 | -0.30844 | 0.22292 | 0.52016 | -0.38056 | 0.01093 | -0.12065 | 0.2528 | -0.05102 | 0.07878 | -0.0338 | -0.04377 | -0.33849 | -0.29025 | 0.22515 |
| | **2'FL** | **3FL** | **DFLac** | **3'SL** | **6'SL** | **LNT** | **LNnT** | **LNFP-I** | **LNFP-II** | **LNFP-III** | **LSTb** | **LSTc** | **DFLNT** | **LNH** | **DSLNT** | **FLNH** | **DFLNH** | **FDSLNH** | **DSLNH** |
| **3FL** | -0.31399 | | 0.09162 | 0.17062 | 0.24231 | 0.24587 | 0.04002 | -0.41964 | 0.77127 | -0.1948 | 0.09899 | -0.05247 | 0.24015 | 0.13788 | 0.05438 | 0.22908 | 0.71503 | 0.78818 | 0.24106 |
| | **2'FL** | **3FL** | **DFLac** | **3'SL** | **6'SL** | **LNT** | **LNnT** | **LNFP-I** | **LNFP-II** | **LNFP-III** | **LSTb** | **LSTc** | **DFLNT** | **LNH** | **DSLNT** | **FLNH** | **DFLNH** | **FDSLNH** | **DSLNH** |
| **DFLac** | 0.14079 | 0.09162 | | 0.61584 | 0.11581 | -0.10533 | 0.14322 | 0.06275 | 0.08334 | 0.09073 | 0.199 | 0.34507 | 0.69882 | -0.07313 | 0.37248 | -0.38665 | 0.06456 | -0.15801 | 0.21775 |
| | **2'FL** | **3FL** | **DFLac** | **3'SL** | **6'SL** | **LNT** | **LNnT** | **LNFP-I** | **LNFP-II** | **LNFP-III** | **LSTb** | **LSTc** | **DFLNT** | **LNH** | **DSLNT** | **FLNH** | **DFLNH** | **FDSLNH** | **DSLNH** |
| **3'SL** | 0.14173 | 0.17062 | 0.61584 | | 0.23105 | 0.05007 | 0.22368 | 0.10943 | 0.23596 | -0.04794 | 0.23052 | 0.32401 | 0.56453 | 0.00462 | 0.41625 | -0.17706 | 0.21074 | -0.01041 | 0.34094 |
| | **2'FL** | **3FL** | **DFLac** | **3'SL** | **6'SL** | **LNT** | **LNnT** | **LNFP-I** | **LNFP-II** | **LNFP-III** | **LSTb** | **LSTc** | **DFLNT** | **LNH** | **DSLNT** | **FLNH** | **DFLNH** | **FDSLNH** | **DSLNH** |
| **6'SL** | 0.01017 | 0.24231 | 0.11581 | 0.23105 | | 0.57762 | 0.24792 | 0.36375 | 0.39508 | 0.13944 | 0.41643 | 0.5909 | 0.30813 | 0.26224 | 0.52578 | 0.37009 | 0.35278 | 0.22481 | 0.66537 |
| | **2'FL** | **3FL** | **DFLac** | **3'SL** | **6'SL** | **LNT** | **LNnT** | **LNFP-I** | **LNFP-II** | **LNFP-III** | **LSTb** | **LSTc** | **DFLNT** | **LNH** | **DSLNT** | **FLNH** | **DFLNH** | **FDSLNH** | **DSLNH** |
| **LNT** | -0.30844 | 0.24587 | -0.10533 | 0.05007 | 0.57762 | | 0.39827 | 0.26597 | 0.58331 | 0.1055 | 0.51413 | 0.22759 | 0.20257 | 0.30939 | 0.55141 | 0.43688 | 0.48781 | 0.28782 | 0.25122 |
| | **2'FL** | **3FL** | **DFLac** | **3'SL** | **6'SL** | **LNT** | **LNnT** | **LNFP-I** | **LNFP-II** | **LNFP-III** | **LSTb** | **LSTc** | **DFLNT** | **LNH** | **DSLNT** | **FLNH** | **DFLNH** | **FDSLNH** | **DSLNH** |
| **LNnT** | 0.22292 | 0.04002 | 0.14322 | 0.22368 | 0.24792 | 0.39827 | | 0.27717 | 0.27714 | 0.05089 | 0.27826 | 0.46085 | 0.35637 | 0.54696 | 0.36535 | 0.2071 | 0.26205 | 0.05766 | 0.20431 |
| | **2'FL** | **3FL** | **DFLac** | **3'SL** | **6'SL** | **LNT** | **LNnT** | **LNFP-I** | **LNFP-II** | **LNFP-III** | **LSTb** | **LSTc** | **DFLNT** | **LNH** | **DSLNT** | **FLNH** | **DFLNH** | **FDSLNH** | **DSLNH** |
| **LNFP-I** | 0.52016 | -0.41964 | 0.06275 | 0.10943 | 0.36375 | 0.26597 | 0.27717 | | -0.24613 | 0.30184 | 0.3009 | 0.41838 | 0.00949 | 0.1306 | 0.41176 | 0.04757 | -0.23515 | -0.37748 | 0.25063 |
| | **2'FL** | **3FL** | **DFLac** | **3'SL** | **6'SL** | **LNT** | **LNnT** | **LNFP-I** | **LNFP-II** | **LNFP-III** | **LSTb** | **LSTc** | **DFLNT** | **LNH** | **DSLNT** | **FLNH** | **DFLNH** | **FDSLNH** | **DSLNH** |
| **LNFP-II** | -0.38056 | 0.77127 | 0.08334 | 0.23596 | 0.39508 | 0.58331 | 0.27714 | -0.24613 | | -0.01369 | 0.43705 | 0.17605 | 0.38695 | 0.20969 | 0.39031 | 0.25472 | 0.88814 | 0.68789 | 0.34406 |
| | **2'FL** | **3FL** | **DFLac** | **3'SL** | **6'SL** | **LNT** | **LNnT** | **LNFP-I** | **LNFP-II** | **LNFP-III** | **LSTb** | **LSTc** | **DFLNT** | **LNH** | **DSLNT** | **FLNH** | **DFLNH** | **FDSLNH** | **DSLNH** |
| **LNFP-III** | 0.01093 | -0.1948 | 0.09073 | -0.04794 | 0.13944 | 0.1055 | 0.05089 | 0.30184 | -0.01369 | | 0.54805 | 0.38534 | 0.12066 | 0.18745 | 0.25419 | 0.06865 | -0.08092 | -0.11589 | 0.10483 |
| | **2'FL** | **3FL** | **DFLac** | **3'SL** | **6'SL** | **LNT** | **LNnT** | **LNFP-I** | **LNFP-II** | **LNFP-III** | **LSTb** | **LSTc** | **DFLNT** | **LNH** | **DSLNT** | **FLNH** | **DFLNH** | **FDSLNH** | **DSLNH** |
| **LSTb** | -0.12065 | 0.09899 | 0.199 | 0.23052 | 0.41643 | 0.51413 | 0.27826 | 0.3009 | 0.43705 | 0.54805 | | 0.46299 | 0.48754 | 0.24075 | 0.69771 | 0.13901 | 0.35714 | 0.09985 | 0.29132 |
| | **2'FL** | **3FL** | **DFLac** | **3'SL** | **6'SL** | **LNT** | **LNnT** | **LNFP-I** | **LNFP-II** | **LNFP-III** | **LSTb** | **LSTc** | **DFLNT** | **LNH** | **DSLNT** | **FLNH** | **DFLNH** | **FDSLNH** | **DSLNH** |
| **LSTc** | 0.2528 | -0.05247 | 0.34507 | 0.32401 | 0.5909 | 0.22759 | 0.46085 | 0.41838 | 0.17605 | 0.38534 | 0.46299 | | 0.50639 | 0.38176 | 0.51352 | 0.05074 | 0.1408 | -0.05992 | 0.5642 |
| | **2'FL** | **3FL** | **DFLac** | **3'SL** | **6'SL** | **LNT** | **LNnT** | **LNFP-I** | **LNFP-II** | **LNFP-III** | **LSTb** | **LSTc** | **DFLNT** | **LNH** | **DSLNT** | **FLNH** | **DFLNH** | **FDSLNH** | **DSLNH** |
| **DFLNT** | -0.05102 | 0.24015 | 0.69882 | 0.56453 | 0.30813 | 0.20257 | 0.35637 | 0.00949 | 0.38695 | 0.12066 | 0.48754 | 0.50639 | | 0.17399 | 0.53217 | -0.20793 | 0.31158 | 0.02663 | 0.2675 |
| | **2'FL** | **3FL** | **DFLac** | **3'SL** | **6'SL** | **LNT** | **LNnT** | **LNFP-I** | **LNFP-II** | **LNFP-III** | **LSTb** | **LSTc** | **DFLNT** | **LNH** | **DSLNT** | **FLNH** | **DFLNH** | **FDSLNH** | **DSLNH** |
| **LNH** | 0.07878 | 0.13788 | -0.07313 | 0.00462 | 0.26224 | 0.30939 | 0.54696 | 0.1306 | 0.20969 | 0.18745 | 0.24075 | 0.38176 | 0.17399 | | 0.21865 | 0.59018 | 0.23766 | 0.34925 | 0.24377 |
| | **2'FL** | **3FL** | **DFLac** | **3'SL** | **6'SL** | **LNT** | **LNnT** | **LNFP-I** | **LNFP-II** | **LNFP-III** | **LSTb** | **LSTc** | **DFLNT** | **LNH** | **DSLNT** | **FLNH** | **DFLNH** | **FDSLNH** | **DSLNH** |
| **DSLNT** | -0.0338 | 0.05438 | 0.37248 | 0.41625 | 0.52578 | 0.55141 | 0.36535 | 0.41176 | 0.39031 | 0.25419 | 0.69771 | 0.51352 | 0.53217 | 0.21865 | | 0.12016 | 0.35829 | -0.0103 | 0.42211 |
| | **2'FL** | **3FL** | **DFLac** | **3'SL** | **6'SL** | **LNT** | **LNnT** | **LNFP-I** | **LNFP-II** | **LNFP-III** | **LSTb** | **LSTc** | **DFLNT** | **LNH** | **DSLNT** | **FLNH** | **DFLNH** | **FDSLNH** | **DSLNH** |
| **FLNH** | -0.04377 | 0.22908 | -0.38665 | -0.17706 | 0.37009 | 0.43688 | 0.2071 | 0.04757 | 0.25472 | 0.06865 | 0.13901 | 0.05074 | -0.20793 | 0.59018 | 0.12016 | | 0.27652 | 0.53493 | 0.3294 |
| | **2'FL** | **3FL** | **DFLac** | **3'SL** | **6'SL** | **LNT** | **LNnT** | **LNFP-I** | **LNFP-II** | **LNFP-III** | **LSTb** | **LSTc** | **DFLNT** | **LNH** | **DSLNT** | **FLNH** | **DFLNH** | **FDSLNH** | **DSLNH** |
| **DFLNH** | -0.33849 | 0.71503 | 0.06456 | 0.21074 | 0.35278 | 0.48781 | 0.26205 | -0.23515 | 0.88814 | -0.08092 | 0.35714 | 0.1408 | 0.31158 | 0.23766 | 0.35829 | 0.27652 | | 0.70102 | 0.38665 |
| | **2'FL** | **3FL** | **DFLac** | **3'SL** | **6'SL** | **LNT** | **LNnT** | **LNFP-I** | **LNFP-II** | **LNFP-III** | **LSTb** | **LSTc** | **DFLNT** | **LNH** | **DSLNT** | **FLNH** | **DFLNH** | **FDSLNH** | **DSLNH** |
| **FDSLNH** | -0.29025 | 0.78818 | -0.15801 | -0.01041 | 0.22481 | 0.28782 | 0.05766 | -0.37748 | 0.68789 | -0.11589 | 0.09985 | -0.05992 | 0.02663 | 0.34925 | -0.0103 | 0.53493 | 0.70102 | | 0.34657 |
| | **2'FL** | **3FL** | **DFLac** | **3'SL** | **6'SL** | **LNT** | **LNnT** | **LNFP-I** | **LNFP-II** | **LNFP-III** | **LSTb** | **LSTc** | **DFLNT** | **LNH** | **DSLNT** | **FLNH** | **DFLNH** | **FDSLNH** | **DSLNH** |
| **DSLNH** | 0.22515 | 0.24106 | 0.21775 | 0.34094 | 0.66537 | 0.25122 | 0.20431 | 0.25063 | 0.34406 | 0.10483 | 0.29132 | 0.5642 | 0.2675 | 0.24377 | 0.42211 | 0.3294 | 0.38665 | 0.34657 | |

**Supplementary Figure S3.** Correlation matrix between all 19 HMOs in secretor children assessed for seroconversion (n=261). Numeric values represent Pearson correlation coefficients.

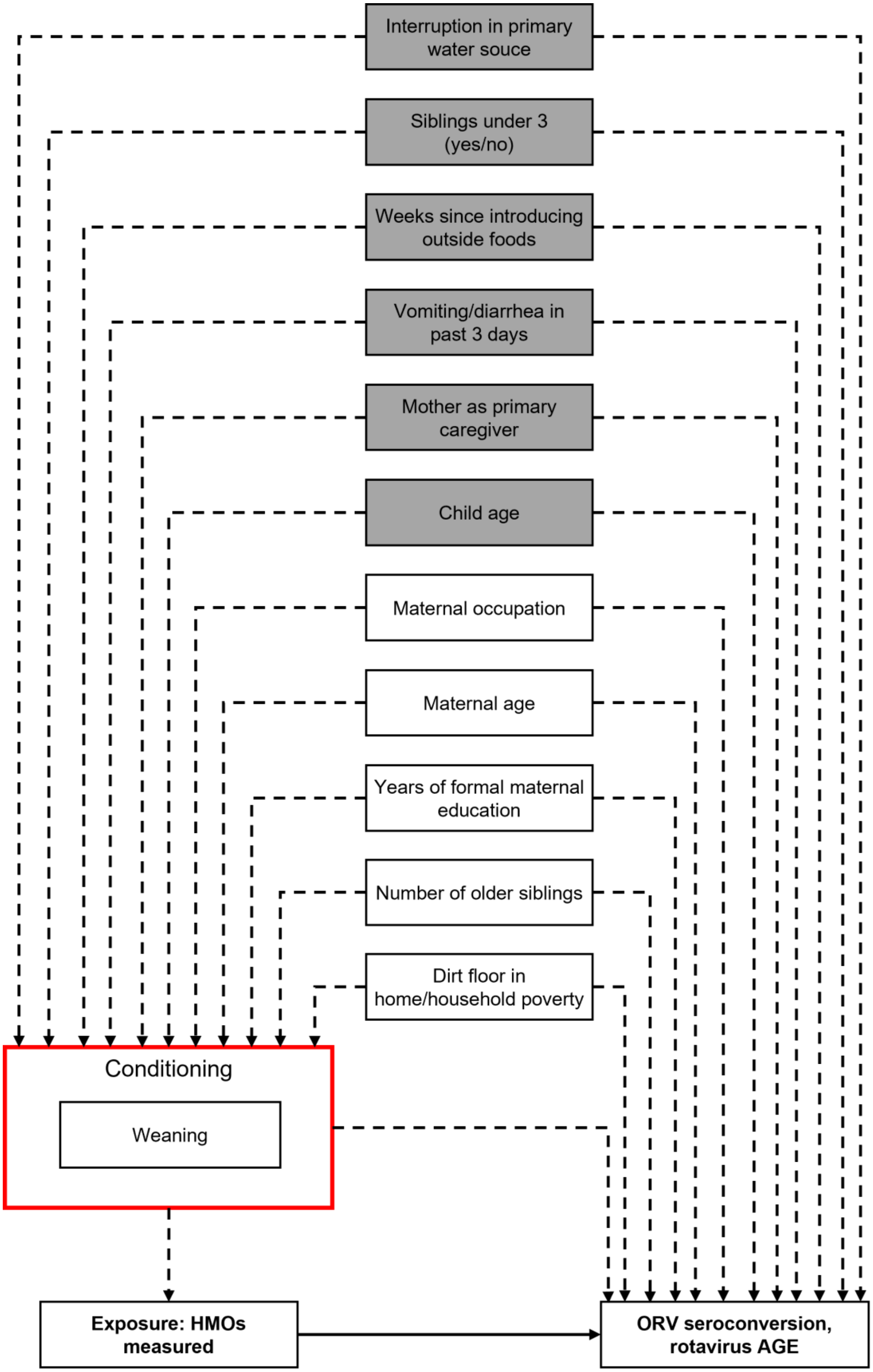
Interruption in primary water souce
Siblings under 3 (yes/no)
Weeks since introducing outside foods
Vomiting/diarrhea in past 3 days
Mother as primary caregiver
Child age
Maternal occupation
Maternal age
Years of formal maternal education
Number of older siblings
Dirt floor in home/household poverty
Conditioning
Weaning
Exposure: HMOs measured
ORV seroconversion, rotavirus AGE

**Supplementary Figure S4:** DAG illustrating the minimally sufficient adjustment set of confounders of the causal relationship between breastfeeding/weaning and ORV seroconversion and 36-month rotavirus AGE incidence outcomes, identified by DAGitty[1]. To correct for selection bias introduced by limiting some analyses to breastfeeding children only, we applied IPCWs to participants for as long as they remained in the risk set (i.e. continued to breastfeed). White boxes represent time-invariant confounders measured at baseline. Gray boxes represent time-varying confounders that were updated at each weekly visit. We ultimately dropped 'weeks since introducing outside foods' from the IPCW calculation as it was highly correlated to child age. Additionally, 'maternal employment' was dropped from the IPCW calculation as we identified potential measurement error and found the variable to be highly correlated to the variable 'mother as primary caregiver'. The final list of confounders included weeks since study entry, children <3 years of age in home (yes/no), primary caregiver to child is mother versus another household member, vomiting or diarrhea in past 3 days, household water supply interrupted in the current month, home dirt floors (approximating household poverty), number of siblings in home, maternal age at child's birth, and years of maternal formal education.

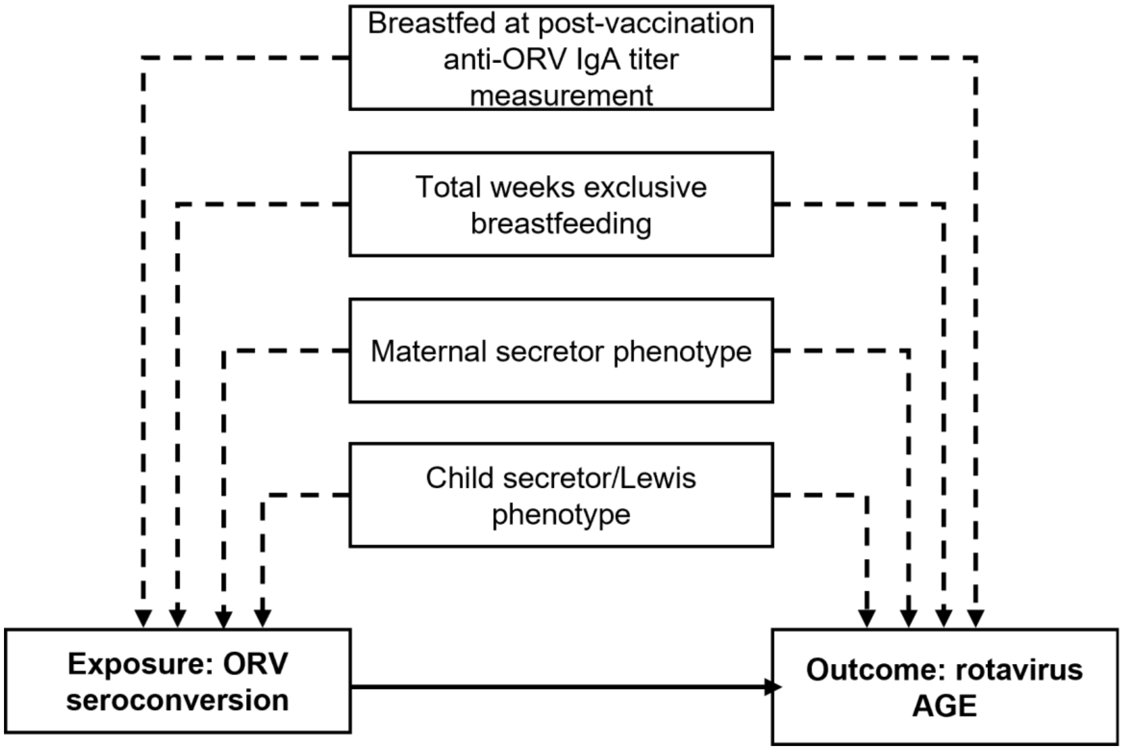


**Supplementary Figure S5:** DAG identifying observed confounders of the causal effect of ORV seroconversion at 1-month after completion of the 2-dose vaccine series on the outcome of 36-month rotavirus AGE incidence. Children were 'breastfed at post-vaccination anti-ORV IgA titer measurement' if their mother reported breastfeeding them the day prior to the closest weekly study visit after the 2nd dose of the ORV.

**Figure S6. Association between human milk oligosaccharide (HMO) concentration and seroconversion (SC) to the monovalent rotavirus vaccine (RV1), crude analysis of all children**

| Comparison | RR (95% CI*) |
|---|---|
| **2'FL** | |
| Middle v. lowest thirds | 1.46 (0.97-2.21) |
| Highest v. lowest thirds | 1.50 (1.00-2.26) |
| **3FL** | |
| Middle v. lowest thirds | 1.03 (0.70-1.51) |
| Highest v. lowest thirds | 1.00 (0.68-1.47) |
| **DFLac** | |
| Middle v. lowest thirds | 0.76 (0.51-1.13) |
| Highest v. lowest thirds | 0.95 (0.66-1.36) |
| **3'SL** | |
| Middle v. lowest thirds | 0.76 (0.51-1.13) |
| Highest v. lowest thirds | 0.95 (0.66-1.36) |
| **6'SL** | |
| Middle v. lowest thirds | 0.97 (0.68-1.39) |
| Highest v. lowest thirds | 0.74 (0.49-1.10) |
| **LNT** | |
| Middle v. lowest thirds | 0.83 (0.58-1.19) |
| Highest v. lowest thirds | 0.68 (0.46-1.01) |
| **LNnT** | |
| Middle v. lowest thirds | 0.92 (0.63-1.33) |
| Highest v. lowest thirds | 0.87 (0.59-1.27) |
| **LNFP-I** | |
| Middle v. lowest thirds | 0.89 (0.61-1.30) |
| Highest v. lowest thirds | 0.89 (0.61-1.30) |
| **LNFP-II** | |
| Middle v. lowest thirds | 0.90 (0.62-1.30) |
| Highest v. lowest thirds | 0.82 (0.56-1.20) |
| **LNFP-III** | |
| Middle v. lowest thirds | 0.89 (0.60-1.31) |
| Highest v. lowest thirds | 0.97 (0.67-1.41) |
| **LSTb** | |
| Middle v. lowest thirds | 0.61 (0.42-0.91) |
| Highest v. lowest thirds | 0.73 (0.51-1.04) |
| **LSTc** | |
| Middle v. lowest thirds | 0.95 (0.67-1.35) |
| Highest v. lowest thirds | 0.69 (0.46-1.04) |
| **DFLNT** | |
| Middle v. lowest thirds | 0.73 (0.51-1.04) |
| Highest v. lowest thirds | 0.61 (0.42-0.91) |
| **LNH** | |
| Middle v. lowest thirds | 0.84 (0.57-1.23) |
| Highest v. lowest thirds | 0.95 (0.65-1.37) |
| **DSLNT** | |
| Middle v. lowest thirds | 0.80 (0.55-1.16) |
| Highest v. lowest thirds | 0.78 (0.53-1.13) |
| **FLNH** | |
| Middle v. lowest thirds | 1.09 (0.74-1.62) |
| Highest v. lowest thirds | 1.13 (0.77-1.66) |
| **DFLNH** | |
| Middle v. lowest thirds | 1.03 (0.71-1.48) |
| Highest v. lowest thirds | 0.83 (0.56-1.24) |
| **FDSLNH** | |
| Middle v. lowest thirds | 1.13 (0.77-1.66) |
| Highest v. lowest thirds | 1.09 (0.74-1.62) |
| **DSLNH** | |
| Middle v. lowest thirds | 1.00 (0.69-1.46) |
| Highest v. lowest thirds | 0.94 (0.64-1.39) |

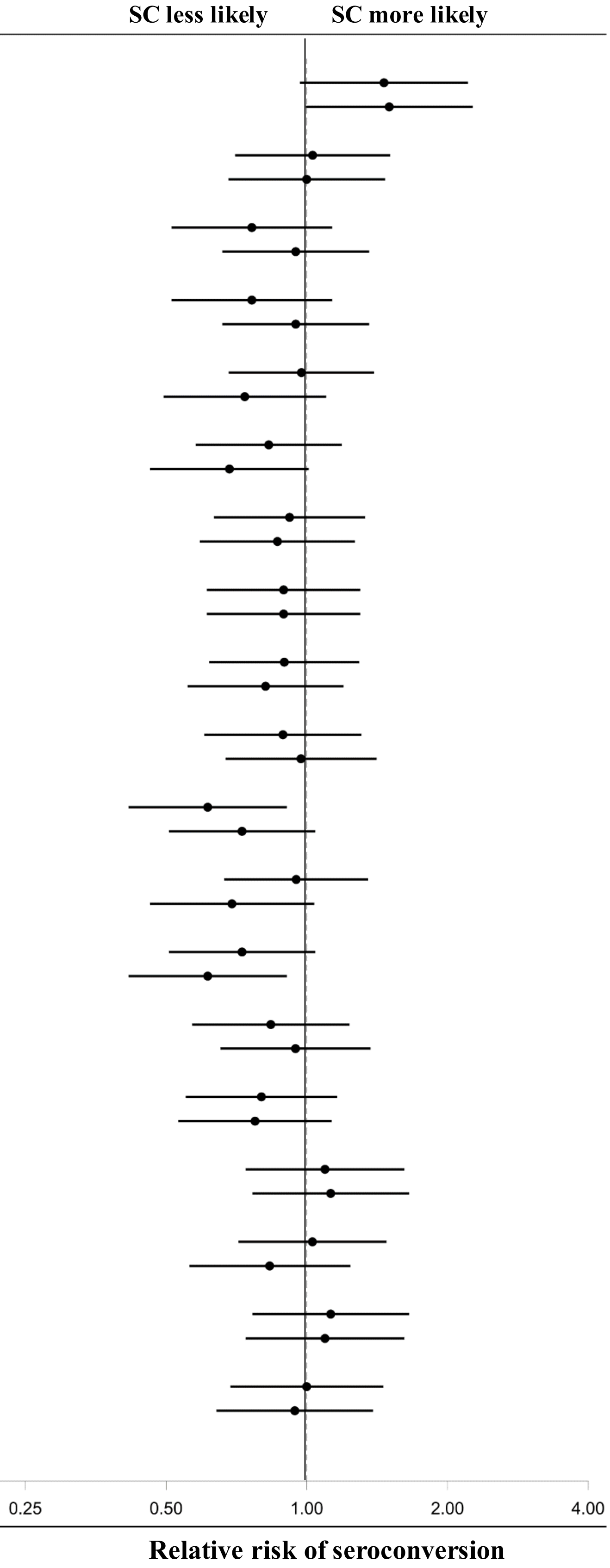

**Supplementary Figure S6:** Unadjusted 5-month risk ratio (RR) and 95% confidence interval (CI) of RV1 seroconversion by maternal HMO concentration among 297 children for whom seroconversion was determined.

**Figure S7. Association between human milk oligosaccharide (HMO) concentration and seroconversion (SC) to the monovalent rotavirus vaccine (RV1), crude analysis among secretor [illegible]**

| Comparison | RR (95% CI*) |
|---|---|
| **2'FL** | |
| Middle v. lowest thirds | 1.46 (0.95-2.23) |
| Highest v. lowest thirds | 1.42 (0.92-2.18) |
| **3FL** | |
| Middle v. lowest thirds | 1.14 (0.76-1.72) |
| Highest v. lowest thirds | 1.18 (0.79-1.77) |
| **DFLac** | |
| Middle v. lowest thirds | 0.77 (0.52-1.16) |
| Highest v. lowest thirds | 0.89 (0.61-1.30) |
| **3'SL** | |
| Middle v. lowest thirds | 0.77 (0.52-1.16) |
| Highest v. lowest thirds | 0.89 (0.61-1.30) |
| **6'SL** | |
| Middle v. lowest thirds | 0.97 (0.67-1.40) |
| Highest v. lowest thirds | 0.69 (0.45-1.05) |
| **LNT** | |
| Middle v. lowest thirds | 0.84 (0.58-1.22) |
| Highest v. lowest thirds | 0.68 (0.45-1.02) |
| **LNnT** | |
| Middle v. lowest thirds | 0.84 (0.57-1.24) |
| Highest v. lowest thirds | 0.85 (0.57-1.26) |
| **LNFP-I** | |
| Middle v. lowest thirds | 0.72 (0.48-1.09) |
| Highest v. lowest thirds | 0.86 (0.59-1.26) |
| **LNFP-II** | |
| Middle v. lowest thirds | 1.07 (0.72-1.57) |
| Highest v. lowest thirds | 0.94 (0.62-1.41) |
| **LNFP-III** | |
| Middle v. lowest thirds | 0.85 (0.57-1.27) |
| Highest v. lowest thirds | 0.97 (0.66-1.43) |
| **LSTb** | |
| Middle v. lowest thirds | 0.61 (0.41-0.91) |
| Highest v. lowest thirds | 0.66 (0.45-0.97) |
| **LSTc** | |
| Middle v. lowest thirds | 0.97 (0.67-1.40) |
| Highest v. lowest thirds | 0.69 (0.45-1.05) |
| **DFLNT** | |
| Middle v. lowest thirds | 0.61 (0.41-0.89) |
| Highest v. lowest thirds | 0.56 (0.37-0.83) |
| **LNH** | |
| Middle v. lowest thirds | 0.80 (0.54-1.19) |
| Highest v. lowest thirds | 0.86 (0.58-1.26) |
| **DSLNT** | |
| Middle v. lowest thirds | 0.76 (0.51-1.12) |
| Highest v. lowest thirds | 0.76 (0.51-1.12) |
| **FLNH** | |
| Middle v. lowest thirds | 1.07 (0.72-1.59) |
| Highest v. lowest thirds | 1.03 (0.69-1.55) |
| **DFLNH** | |
| Middle v. lowest thirds | 1.03 (0.70-1.53) |
| Highest v. lowest thirds | 0.97 (0.65-1.45) |
| **FDSLNH** | |
| Middle v. lowest thirds | 1.26 (0.84-1.90) |
| Highest v. lowest thirds | 1.19 (0.78-1.80) |
| **DSLNH** | |
| Middle v. lowest thirds | 1.24 (0.84-1.83) |
| Highest v. lowest thirds | 0.97 (0.63-1.48) |

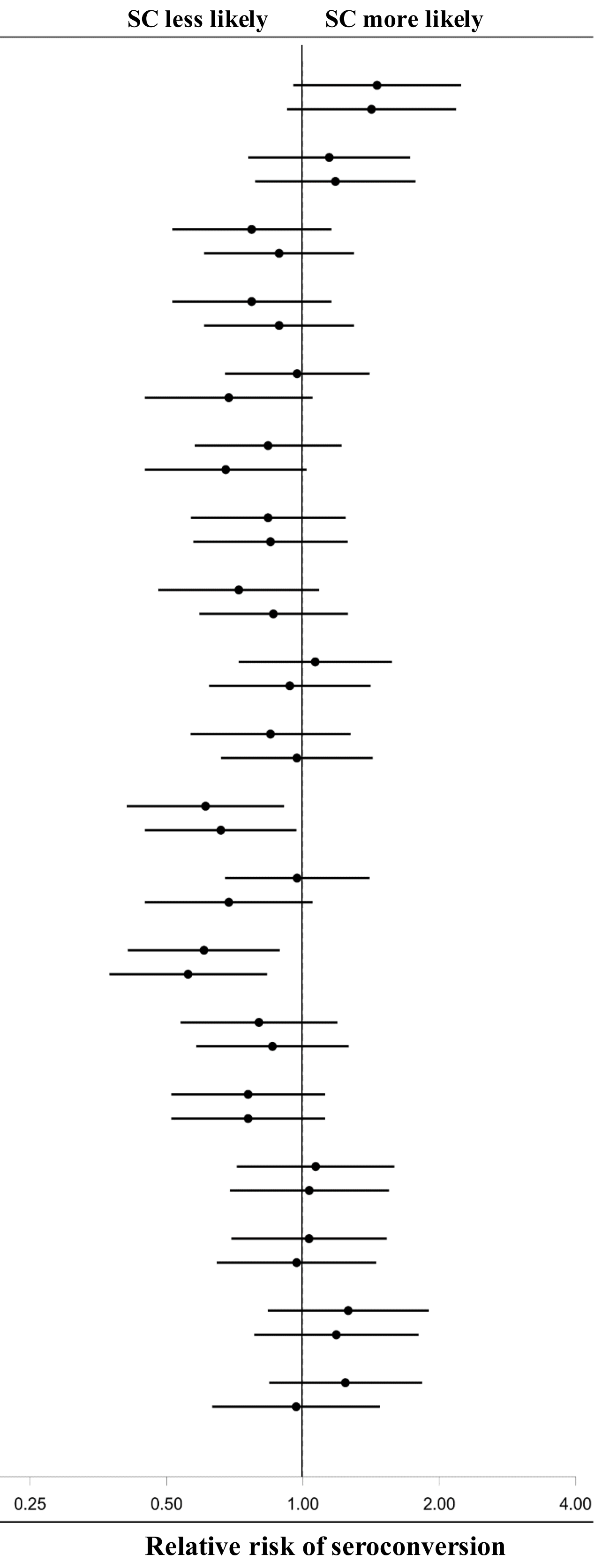

**Supplementary Figure S7:** Unadjusted 5-month risk ratio (RR) and 95% confidence interval (CI) of RV1 seroconversion by maternal HMO concentration among 246 secretor children for whom seroconversion was determined.

**Figure S8. Association between human milk oligosaccharide (HMO) concentration and rotavirus acute gastroenteritis (AGE), crude analysis of all children**

| Comparison | RD (95% CI*) |
|---|---|
| **2'FL** | |
| Middle v. lowest thirds | 0.054 (-0.052- 0.161) |
| Highest v. lowest thirds | 0.104 (-0.007- 0.215) |
| **3FL** | |
| Middle v. lowest thirds | 0.002 (-0.107- 0.111) |
| Highest v. lowest thirds | 0.047 (-0.068- 0.162) |
| **DFLac** | |
| Middle v. lowest thirds | 0.006 (-0.110- 0.122) |
| Highest v. lowest thirds | -0.023 (-0.134- 0.089) |
| **3'SL** | |
| Middle v. lowest thirds | 0.015 (-0.098- 0.128) |
| Highest v. lowest thirds | 0.005 (-0.108- 0.117) |
| **6'SL** | |
| Middle v. lowest thirds | 0.087 (-0.029- 0.204) |
| Highest v. lowest thirds | 0.001 (-0.104- 0.107) |
| **LNT** | |
| Middle v. lowest thirds | -0.013 (-0.115- 0.089) |
| Highest v. lowest thirds | 0.119 ( 0.004- 0.234) |
| **LNnT** | |
| Middle v. lowest thirds | 0.009 (-0.102- 0.120) |
| Highest v. lowest thirds | 0.034 (-0.080- 0.149) |
| **LNFP-I** | |
| Middle v. lowest thirds | 0.131 ( 0.026- 0.237) |
| Highest v. lowest thirds | 0.135 ( 0.028- 0.241) |
| **LNFP-II** | |
| Middle v. lowest thirds | -0.057 (-0.162- 0.048) |
| Highest v. lowest thirds | 0.058 (-0.061- 0.177) |
| **LNFP-III** | |
| Middle v. lowest thirds | -0.002 (-0.115- 0.110) |
| Highest v. lowest thirds | 0.008 (-0.106- 0.122) |
| **LSTb** | |
| Middle v. lowest thirds | 0.075 (-0.039- 0.189) |
| Highest v. lowest thirds | 0.030 (-0.077- 0.137) |
| **LSTc** | |
| Middle v. lowest thirds | -0.048 (-0.158- 0.061) |
| Highest v. lowest thirds | 0.016 (-0.104- 0.136) |
| **DFLNT** | |
| Middle v. lowest thirds | -0.037 (-0.150- 0.076) |
| Highest v. lowest thirds | -0.009 (-0.124- 0.106) |
| **LNH** | |
| Middle v. lowest thirds | 0.031 (-0.080- 0.142) |
| Highest v. lowest thirds | 0.055 (-0.057- 0.167) |
| **DSLNT** | |
| Middle v. lowest thirds | 0.112 (-0.001- 0.225) |
| Highest v. lowest thirds | 0.052 (-0.053- 0.158) |
| **FLNH** | |
| Middle v. lowest thirds | 0.026 (-0.082- 0.134) |
| Highest v. lowest thirds | 0.069 (-0.045- 0.183) |
| **DFLNH** | |
| Middle v. lowest thirds | 0.012 (-0.098- 0.122) |
| Highest v. lowest thirds | 0.050 (-0.064- 0.163) |
| **FDSLNH** | |
| Middle v. lowest thirds | -0.013 (-0.124- 0.097) |
| Highest v. lowest thirds | 0.027 (-0.087- 0.141) |
| **DSLNH** | |
| Middle v. lowest thirds | 0.056 (-0.055- 0.167) |
| Highest v. lowest thirds | 0.036 (-0.075- 0.146) |

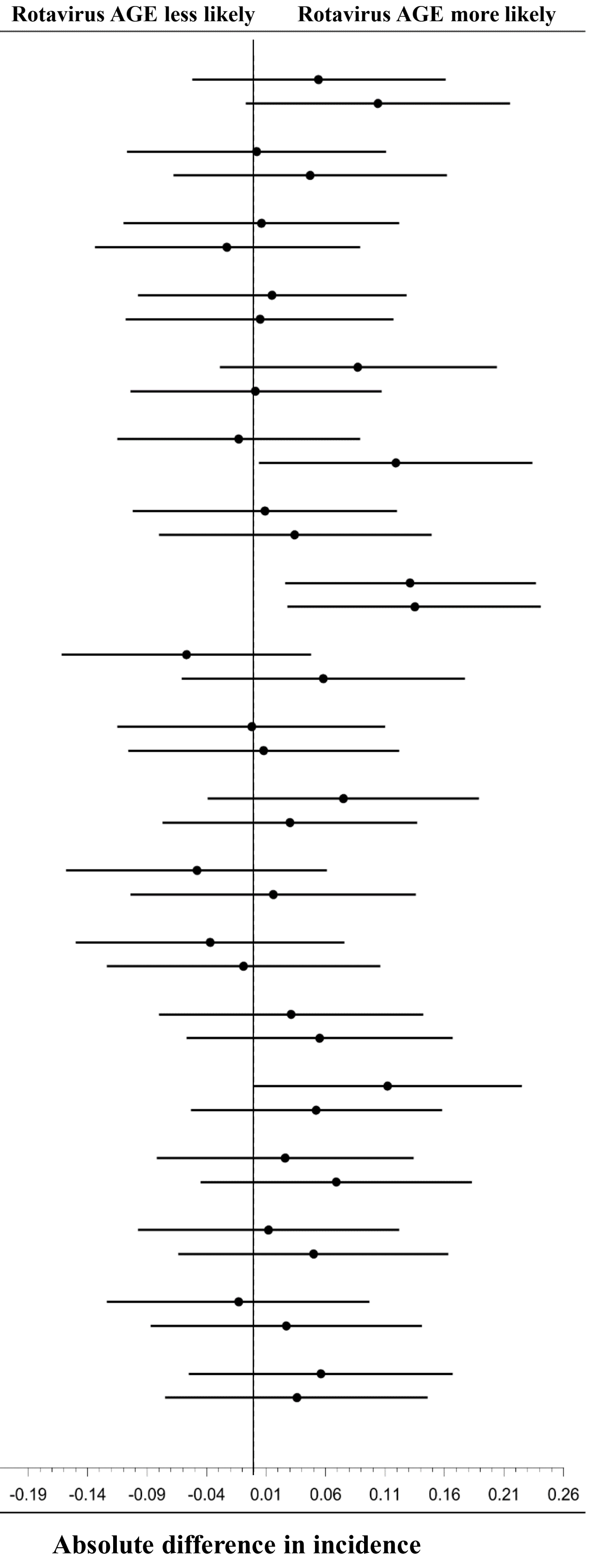

**Supplementary Figure S8:** Crude 36-month risk difference (RD) and 95% CI of rotavirus AGE by maternal HMO composition among 282 cohort children with HMOs characterized who remained in the study ≥ 1- month post-vaccination and who had seroconversion analyzed.

### Figure S9. Association between human milk oligosaccharide (HMO) concentration and rotavirus acute gastroenteritis (AGE), crude analysis of secretor children

| Comparison | RD (95% CI*) |
|---|---|
| **2'FL** | |
| Middle v. lowest thirds | 0.047 (-0.073- 0.167) |
| Highest v. lowest thirds | 0.093 (-0.031- 0.218) |
| **3FL** | |
| Middle v. lowest thirds | -0.013 (-0.135- 0.110) |
| Highest v. lowest thirds | 0.030 (-0.098- 0.159) |
| **DFLac** | |
| Middle v. lowest thirds | -0.075 (-0.203- 0.053) |
| Highest v. lowest thirds | -0.066 (-0.195- 0.062) |
| **3'SL** | |
| Middle v. lowest thirds | 0.032 (-0.096- 0.159) |
| Highest v. lowest thirds | -0.008 (-0.131- 0.115) |
| **6'SL** | |
| Middle v. lowest thirds | 0.103 (-0.027- 0.232) |
| Highest v. lowest thirds | 0.002 (-0.115- 0.118) |
| **LNT** | |
| Middle v. lowest thirds | -0.030 (-0.144- 0.083) |
| Highest v. lowest thirds | 0.119 (-0.011- 0.249) |
| **LNnT** | |
| Middle v. lowest thirds | 0.007 (-0.120- 0.134) |
| Highest v. lowest thirds | -0.003 (-0.130- 0.123) |
| **LNFP-I** | |
| Middle v. lowest thirds | 0.080 (-0.040- 0.200) |
| Highest v. lowest thirds | 0.099 (-0.023- 0.222) |
| **LNFP-II** | |
| Middle v. lowest thirds | -0.066 (-0.181- 0.050) |
| Highest v. lowest thirds | 0.068 (-0.064- 0.201) |
| **LNFP-III** | |
| Middle v. lowest thirds | 0.020 (-0.105- 0.146) |
| Highest v. lowest thirds | 0.020 (-0.105- 0.146) |
| **LSTb** | |
| Middle v. lowest thirds | 0.102 (-0.023- 0.227) |
| Highest v. lowest thirds | 0.061 (-0.057- 0.180) |
| **LSTc** | |
| Middle v. lowest thirds | -0.092 (-0.212- 0.028) |
| Highest v. lowest thirds | 0.015 (-0.121- 0.151) |
| **DFLNT** | |
| Middle v. lowest thirds | -0.082 (-0.206- 0.041) |
| Highest v. lowest thirds | -0.013 (-0.143- 0.118) |
| **LNH** | |
| Middle v. lowest thirds | 0.011 (-0.116- 0.138) |
| Highest v. lowest thirds | 0.010 (-0.114- 0.135) |
| **DSLNT** | |
| Middle v. lowest thirds | 0.093 (-0.031- 0.218) |
| Highest v. lowest thirds | 0.048 (-0.072- 0.169) |
| **FLNH** | |
| Middle v. lowest thirds | 0.017 (-0.105- 0.138) |
| Highest v. lowest thirds | 0.054 (-0.074- 0.181) |
| **DFLNH** | |
| Middle v. lowest thirds | 0.001 (-0.123- 0.125) |
| Highest v. lowest thirds | 0.031 (-0.095- 0.157) |
| **FDSLNH** | |
| Middle v. lowest thirds | 0.016 (-0.111- 0.142) |
| Highest v. lowest thirds | 0.004 (-0.119- 0.128) |
| **DSLNH** | |
| Middle v. lowest thirds | 0.048 (-0.073- 0.169) |
| Highest v. lowest thirds | 0.056 (-0.070- 0.181) |

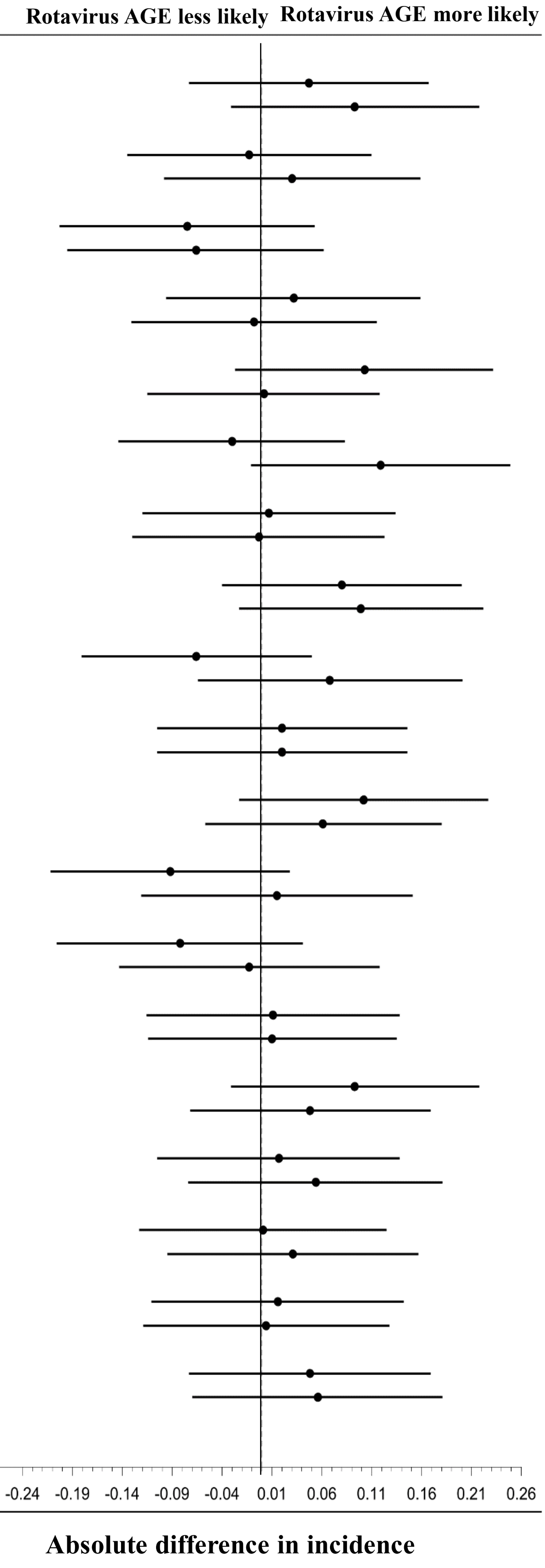

**Supplementary Figure S9:** Crude 36-month risk difference (RD) and 95% CI of wild-type rotavirus AGE by maternal HMO composition among 246 secretor cohort children with HMOs characterized who had remained in the study ≥ 1-month post-vaccination and who had been assessed for seroconversion.

**Figure S10. Association between human milk oligosaccharide (HMO) concentration and rotavirus acute gastroenteritis (AGE), adjusted analysis of all children, censored after weaning**

| Comparison | RD (95% CI*) |
|---|---|
| **2'FL** | |
| Middle v. lowest thirds | -0.009 (-0.140- 0.122) |
| Highest v. lowest thirds | 0.009 (-0.122- 0.140) |
| **3FL** | |
| Middle v. lowest thirds | -0.120 (-0.355- 0.115) |
| Highest v. lowest thirds | -0.011 (-0.300- 0.278) |
| **DFLac** | |
| Middle v. lowest thirds | 0.023 (-0.086- 0.132) |
| Highest v. lowest thirds | -0.032 (-0.132- 0.067) |
| **3'SL** | |
| Middle v. lowest thirds | 0.026 (-0.127- 0.180) |
| Highest v. lowest thirds | -0.033 (-0.172- 0.106) |
| **6'SL** | |
| Middle v. lowest thirds | 0.157 (-0.164- 0.477) |
| Highest v. lowest thirds | -0.147 (-0.440- 0.147) |
| **LNT** | |
| Middle v. lowest thirds | -0.034 (-0.173- 0.105) |
| Highest v. lowest thirds | 0.335 ( 0.094- 0.576) |
| **LNnT** | |
| Middle v. lowest thirds | -0.118 (-0.389- 0.154) |
| Highest v. lowest thirds | -0.104 (-0.376- 0.167) |
| **LNFP-I** | |
| Middle v. lowest thirds | 0.149 ( 0.036- 0.261) |
| Highest v. lowest thirds | 0.194 ( 0.032- 0.356) |
| **LNFP-II** | |
| Middle v. lowest thirds | 0.109 (-0.148- 0.367) |
| Highest v. lowest thirds | 0.129 (-0.041- 0.298) |
| **LNFP-III** | |
| Middle v. lowest thirds | -0.003 (-0.187- 0.181) |
| Highest v. lowest thirds | -0.095 (-0.255- 0.066) |
| **LSTb** | |
| Middle v. lowest thirds | 0.014 (-0.232- 0.259) |
| Highest v. lowest thirds | -0.071 (-0.289- 0.147) |
| **LSTc** | |
| Middle v. lowest thirds | 0.046 (-0.125- 0.218) |
| Highest v. lowest thirds | 0.001 (-0.166- 0.169) |
| **DFLNT** | |
| Middle v. lowest thirds | -0.071 (-0.292- 0.151) |
| Highest v. lowest thirds | -0.010 (-0.259- 0.240) |
| **LNH** | |
| Middle v. lowest thirds | 0.012 (-0.115- 0.139) |
| Highest v. lowest thirds | 0.083 (-0.073- 0.239) |
| **DSLNT** | |
| Middle v. lowest thirds | 0.093 (-0.084- 0.270) |
| Highest v. lowest thirds | 0.022 (-0.144- 0.189) |
| **FLNH** | |
| Middle v. lowest thirds | 0.067 (-0.080- 0.213) |
| Highest v. lowest thirds | 0.096 (-0.043- 0.235) |
| **DFLNH** | |
| Middle v. lowest thirds | -0.078 (-0.294- 0.137) |
| Highest v. lowest thirds | -0.120 (-0.321- 0.080) |
| **FDSLNH** | |
| Middle v. lowest thirds | -0.088 (-0.204- 0.028) |
| Highest v. lowest thirds | -0.020 (-0.145- 0.105) |
| **DSLNH** | |
| Middle v. lowest thirds | 0.085 (-0.042- 0.211) |
| Highest v. lowest thirds | 0.070 (-0.093- 0.233) |

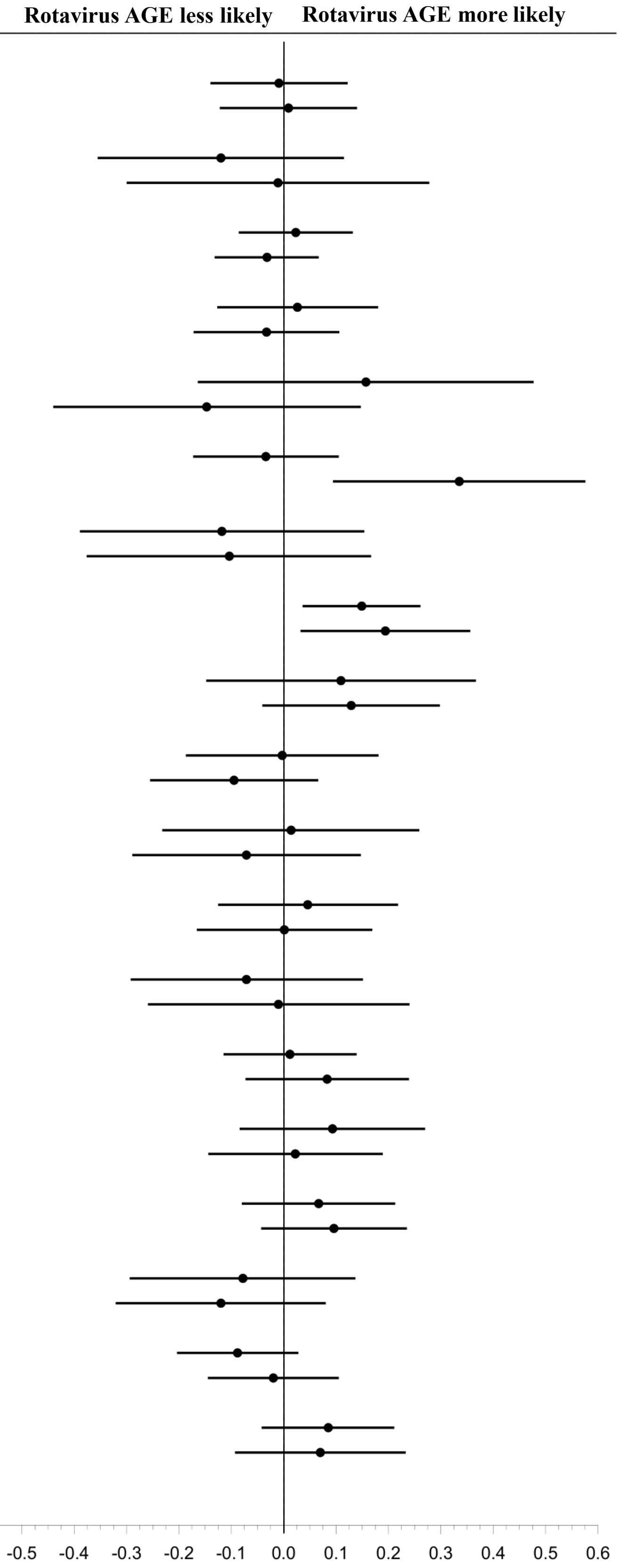


**Absolute difference in incidence proportion**

**Figure S10:** Adjusted 36-month RD and 95% CI of rotavirus AGE by maternal HMO composition among all 282 cohort children with HMOs characterized, who remained in the study ≥ 1 month post-vaccination, and who had seroconversion analyzed. Follow-up was limited to children who had breastfed within the previous 2 weeks. In addition to IPTWs applied to account for confounding bias, we also weighted the remaining children with the inverse probability of censoring (IPCWs) to account for selection bias induced by censoring children after weaning.

**Figure S11. Association between human milk oligosaccharide (HMO) concentration and rotavirus acute gastroenteritis (AGE), adjusted analysis of secretor children, censored after weaning**

| Comparison | RD (95% CI*) |
|---|---|
| **2'FL** | |
| Middle v. lowest thirds | 0.003 (-0.123- 0.128) |
| Highest v. lowest thirds | 0.030 (-0.100- 0.160) |
| **3FL** | |
| Middle v. lowest thirds | -0.179 (-0.446- 0.088) |
| Highest v. lowest thirds | -0.117 (-0.405- 0.171) |
| **DFLac** | |
| Middle v. lowest thirds | 0.083 (-0.152- 0.318) |
| Highest v. lowest thirds | -0.040 (-0.180- 0.101) |
| **3'SL** | |
| Middle v. lowest thirds | 0.063 (-0.114- 0.240) |
| Highest v. lowest thirds | -0.007 (-0.187- 0.173) |
| **6'SL** | |
| Middle v. lowest thirds | 0.224 (-0.120- 0.568) |
| Highest v. lowest thirds | -0.154 (-0.458- 0.149) |
| **LNT** | |
| Middle v. lowest thirds | -0.004 (-0.147- 0.139) |
| Highest v. lowest thirds | 0.294 ( 0.100- 0.488) |
| **LNnT** | |
| Middle v. lowest thirds | 0.004 (-0.192- 0.199) |
| Highest v. lowest thirds | -0.006 (-0.192- 0.181) |
| **LNFP-I** | |
| Middle v. lowest thirds | 0.172 ( 0.046- 0.298) |
| Highest v. lowest thirds | 0.139 ( 0.029- 0.250) |
| **LNFP-II** | |
| Middle v. lowest thirds | 0.004 (-0.160- 0.168) |
| Highest v. lowest thirds | 0.126 (-0.036- 0.288) |
| **LNFP-III** | |
| Middle v. lowest thirds | 0.029 (-0.144- 0.201) |
| Highest v. lowest thirds | -0.062 (-0.221- 0.097) |
| **LSTb** | |
| Middle v. lowest thirds | -0.026 (-0.305- 0.253) |
| Highest v. lowest thirds | -0.008 (-0.317- 0.301) |
| **LSTc** | |
| Middle v. lowest thirds | 0.032 (-0.150- 0.214) |
| Highest v. lowest thirds | 0.016 (-0.166- 0.198) |
| **DFLNT** | |
| Middle v. lowest thirds | 0.017 (-0.099- 0.133) |
| Highest v. lowest thirds | 0.154 (-0.047- 0.354) |
| **LNH** | |
| Middle v. lowest thirds | -0.004 (-0.150- 0.143) |
| Highest v. lowest thirds | 0.078 (-0.083- 0.240) |
| **DSLNT** | |
| Middle v. lowest thirds | 0.172 ( 0.012- 0.333) |
| Highest v. lowest thirds | 0.135 (-0.027- 0.297) |
| **FLNH** | |
| Middle v. lowest thirds | 0.075 (-0.067- 0.216) |
| Highest v. lowest thirds | 0.100 (-0.038- 0.237) |
| **DFLNH** | |
| Middle v. lowest thirds | -0.104 (-0.343- 0.134) |
| Highest v. lowest thirds | -0.137 (-0.363- 0.089) |
| **FDSLNH** | |
| Middle v. lowest thirds | -0.086 (-0.215- 0.043) |
| Highest v. lowest thirds | 0.012 (-0.141- 0.165) |
| **DSLNH** | |
| Middle v. lowest thirds | 0.092 (-0.060- 0.244) |
| Highest v. lowest thirds | 0.108 (-0.078- 0.295) |

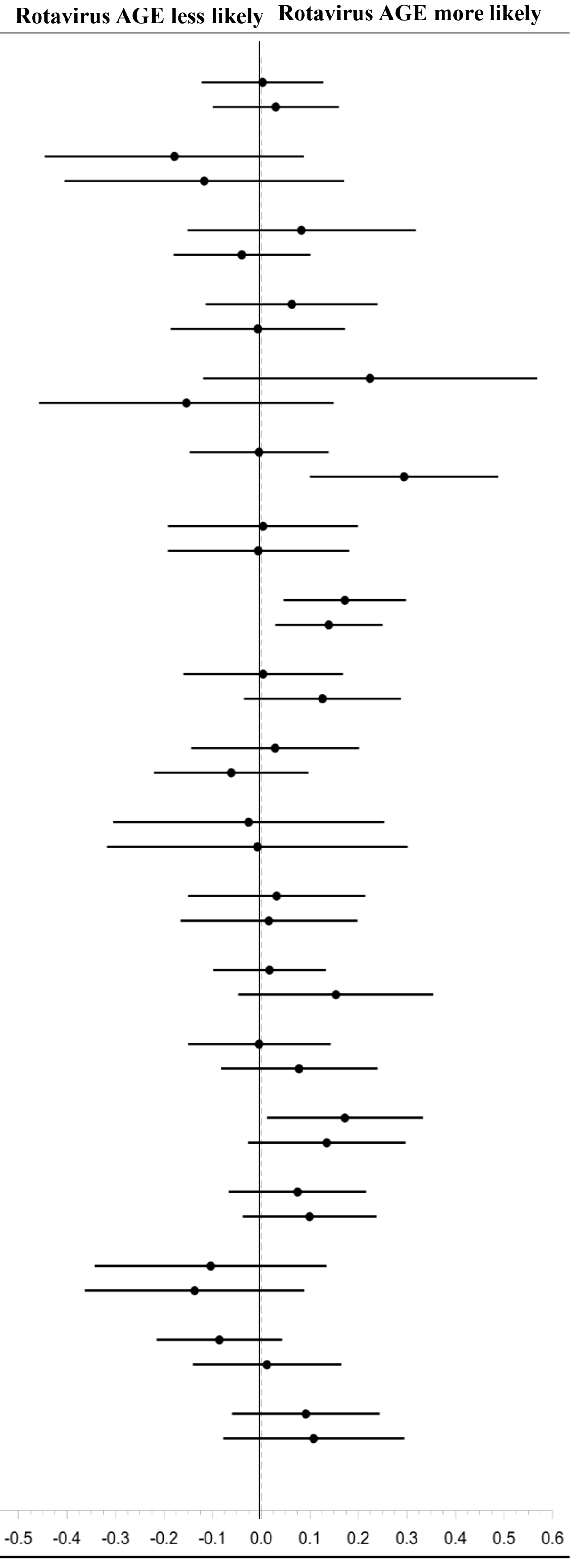


**Absolute difference in incidence**

**Supplementary Figure S11:** Adjusted 36-month risk difference (RD) and 95% CI of rotavirus AGE by maternal HMO composition among 246 secretor cohort children with HMOs characterized who had remained in the study ≥ 1 month post-vaccination and who had been assessed for seroconversion. Follow-up was limited to children who had breastfed within the previous 2 weeks. In addition to IPTWs applied to account for confounding bias, we also weighted the remaining children with the inverse probability of censoring (IPCWs) to account for selection bias induced by censoring children after weaning.
